# Prescribing of antipsychotics for people diagnosed with severe mental illness in UK primary care: A 20-year investigation of who receives treatment, with which agents, and at what doses

**DOI:** 10.1101/2024.03.26.24304727

**Authors:** Alvin Richards-Belle, Naomi Launders, Sarah Hardoon, Kenneth K.C. Man, Elvira Bramon, David P.J. Osborn, Joseph F. Hayes

## Abstract

**Background:** Contemporary data relating to antipsychotic prescribing in UK primary care for patients diagnosed with severe mental illness (SMI) are lacking.

**Aims:** To describe contemporary patterns of antipsychotic prescribing in UK primary care for patients diagnosed with SMI.

**Methods:** Cohort study of patients with an SMI diagnosis (i.e., schizophrenia, bipolar disorder, other non-organic psychoses) first recorded in primary care between 2000-2017 derived from Clinical Practice Research Datalink. Patients were considered exposed to antipsychotics if prescribed at least one antipsychotic in primary care between 2000-2019. We compared characteristics of patients prescribed and not prescribed antipsychotics; calculated annual prevalence rates for antipsychotic prescribing; and computed average daily antipsychotic doses stratified by patient characteristics.

**Results:** Of 309,378 patients first diagnosed with an SMI in primary care between 2000-2017, 212,618 (68.7%) were prescribed an antipsychotic between 2000-2019. Antipsychotic prescribing prevalence was 426 (95% CI, 420-433) per 1,000 patients in the year 2000, reaching a peak of 550 (547-553) in 2016, decreasing to 470 (468-473) in 2019. The proportion prescribed antipsychotics was higher amongst patients diagnosed with schizophrenia (81.0%) than with bipolar disorder (64.6%) and other non-organic psychoses (65.7%). Olanzapine, quetiapine, risperidone, and aripiprazole accounted for 78.8% of all prescriptions. Higher mean olanzapine equivalent total daily doses were prescribed to patients with the following characteristics: schizophrenia diagnosis, ethnic minority status, male sex, younger age, and greater deprivation.

**Conclusions:** Antipsychotic prescribing is dominated by olanzapine, quetiapine, risperidone, and aripiprazole. Two thirds of patients with diagnosed SMI were prescribed antipsychotics in primary care, but this proportion varied according to SMI diagnosis. There were disparities in both receipt and dose of antipsychotics across subgroups - further efforts are needed to understand why certain groups are prescribed higher doses and whether they require dose optimisation to minimise side effects.

## INTRODUCTION

Antipsychotic medications are primarily indicated for the management of psychotic symptoms associated with severe mental illnesses (SMI), such as schizophrenia and bipolar disorder, and were prescribed to 810,000 patients in England alone in 2022/23 (an increase of 22% from 2015/16).^1^ Amongst patients diagnosed with schizophrenia-spectrum disorders, antipsychotic use (versus non-use) is associated with a significantly lower long-term mortality rate.^2^ Despite this overall benefit, antipsychotic agents vary in their propensity for adverse reactions - with cardiometabolic effects, such as weight gain, dyslipidaemia, and hyperglycaemia, major concerns of “second-generation” antipsychotics, such as olanzapine, quetiapine, and risperidone.^3^

In the United Kingdom (UK), primary care services are responsible for the long-term prescribing of antipsychotics to patients diagnosed with SMI. Data relating to antipsychotic prescribing in primary care are therefore essential for monitoring trends and identifying priorities for quality improvement and research. However, contemporary data on the SMI population are limited. Most recent reports have focussed on other diagnoses, such as dementia^4^ or personality disorders,^5^ or on all-cause prescribing in children and young people^6^ and adults.^7^

Earlier studies have documented antipsychotic prescribing practice in primary care for patients diagnosed with SMI.^8,9^ Prah *et al.* investigated trends in schizophrenia, 1998-2007,^8^ and Hayes *et al.* investigated bipolar disorder, 1995-2009.^9^ Both studies (1) illustrated the shift from prescribing first- to second-generation antipsychotics, (2) highlighted olanzapine, risperidone, and quetiapine as the most frequently prescribed antipsychotics, and (3) documented increases in the proportion of time spent receiving antipsychotic treatment, particularly for women (those aged ≥45 diagnosed with schizophrenia^8^ and those 18-30 diagnosed with bipolar disorder^9^). A more recent study reported further increases in antipsychotic prescribing to patients diagnosed with bipolar disorder (from 37% of patients in 2001 to 45% by 2018), with quetiapine, olanzapine, and aripiprazole now the most frequently prescribed.^10^ Whether prescribing for schizophrenia and other psychoses has followed these trends is unknown, particularly following the licensing of aripiprazole in 2004.^11^

Several studies report disparities in aspects of antipsychotic prescribing in UK primary care. A 2006 study, in one London borough, compared the primary care management of Black versus White patients diagnosed with psychosis and reported that Black patients had greater odds of being prescribed long-acting injectable antipsychotics.^12^ A study (2005-2015) of diverse psychiatric diagnoses identified that men were, on average, prescribed higher antipsychotic doses than women - but results were not stratified by SMI diagnosis.^13^ Further contemporary exploration of these and other potential disparities, including stratification by age and deprivation, are needed in order to inform efforts to achieve equity of care.

### Aim and objectives

In order to inform future quality improvement and research into the safer prescribing of antipsychotics, the overall aim of this study was to describe contemporary (2000-2019) patterns of antipsychotic prescribing for people diagnosed with SMI in UK primary care. Specific objectives were:

1. to compare the characteristics of patients diagnosed with SMI prescribed and not prescribed antipsychotics in primary care;
2. to describe the most frequently prescribed antipsychotics in primary care and how this may have changed over time; and
3. to describe the average prescribed daily antipsychotic dose over the first year of prescribing and explore whether doses vary according to diagnosis, ethnicity, age, sex, and deprivation.

## METHODS

### Study design and data source

We conducted a longitudinal cohort study, using data from Clinical Practice Research Datalink (CPRD), to investigate antipsychotic prescribing from 1 January 2000 to 31 December 2019 in a cohort of people first diagnosed with SMI in primary care between 1 January 2000 and 31 December 2017. The study design is summarised in Supplementary Figure 1.

CPRD encompasses two databases (Aurum^14^ and GOLD^15^) which, collectively, contain the de-identified primary care records of over 62 million (current and historic) patients from participating National Health Service primary care practices. Over 98% of the UK population are registered in primary care and CPRD has been shown to be broadly representative, with coverage of almost a quarter of the current population.^16,17^ CPRD contains coded information on consultations, prescriptions, observations, and referrals. We used data from the May 2022 and April 2023 builds of Aurum and GOLD, respectively.

#### Ethics and consent

All procedures involving patients were approved by the East Midlands - Derby Research Ethics Committee (reference: 21/EM/0265). This study was reviewed by the Independent Scientific Advisory Committee of CPRD (protocol no. 21_000729). All data sent by GP practices to CPRD are anonymised and therefore individual patient consent was not required (patients are able to opt-out from their data being shared).

### Participants

The cohort comprised patients actively registered in primary care between 2000-2019 identified as first receiving an SMI diagnosis in their primary care record between 2000-2017. SMI diagnosis was defined as a recorded Read or EMIS® code indicating diagnosis of schizophrenia, bipolar disorder, or other non-organic psychoses (e.g., psychotic episodes, schizoaffective disorders, delusional disorder, non-organic psychosis not otherwise specified) (see the online repository for the code list, which was verified by a clinician (JFH)). SMI diagnoses are typically made by psychiatrists in secondary care and subsequently communicated to primary care. The validity of SMI diagnoses recorded in primary care has been established.^18^

### Outcomes: Antipsychotics

Patients were considered exposed to antipsychotics if prescribed at least one antipsychotic in primary care during the study period (2000-2019). Antipsychotics could be initiated by general practitioners or specialists (e.g. psychiatrists), but must have been issued through primary care (standard practice for longer-term community prescriptions in the UK).^19^ Unless otherwise specified, antipsychotic prescription could pre-date the recording of SMI diagnosis in primary care (provided it was within the study period), given that antipsychotics may be initiated before a specific SMI diagnosis is formulated and/or communicated to primary care. Whilst antipsychotic prescription could pre-date SMI diagnosis, the requirement for first-recorded SMI diagnosis between 2000-2017 allowed for each patient to accrue at least up to two years follow-up post-diagnosis (assuming they remained alive and registered in primary care).

#### Prescriptions of antipsychotics (objective 1 and 2)

Antipsychotic medications (current and withdrawn) were identified through review of national and international reference sources.^20,21^ Search strategies, based on antipsychotic generic and common brand names (Supplementary Table 1), were developed to identify relevant product codes in CPRD code dictionaries. For patients in the cohort, product codes were then used to extract data from prescription records, including product name, ingredient, prescription issue date, strength, formulation, route of administration, quantity, duration, and de-identified free-text containing dosing instructions. We included both oral and injectable antipsychotics, but did not include prochlorperazine as an antipsychotic given it is primarily used as an antiemetic.

#### Antipsychotic dose (objective 3)

We derived the total daily prescribed oral antipsychotic dose for each of (up to) the first 12 prescription dates for patients newly initiating antipsychotics in the study period (i.e., where patients had no identified prescriptions for antipsychotics in primary care prior to the study period). We considered doses of all tablet (e.g., extended release, sublingual) and liquid, but not injectable, antipsychotic formulations. Free-text dosage instructions (e.g., “take five tablets per day”) were converted to numerical quantities using a text-mining algorithm implemented in the R package *doseminer*.^22^ To enable comparison across agents, calculated doses were then converted to olanzapine equivalents according to the Defined Daily Dose (DDD) method^23^ using *chlorpromazineR*^24^ (cariprazine and droperidol were not reported in the DDD method,^23^ equivalence formulae for these antipsychotics came from references ^25^ and ^26^, respectively). In the case of multiple prescriptions issued on the same date, we considered up to three unique prescriptions of each antipsychotic prescribed on a given date (>3 unique prescriptions of one medication was considered potentially erroneous).

### Stratifying variables

We extracted additional variables from CPRD, to characterise the cohort and for stratified analyses. These included: year of birth, sex, ethnicity, geographic region, relative deprivation, date of first SMI diagnosis, SMI diagnosis, and prescriptions of antidepressants and mood stabilisers. Where a patient had multiple ethnicity categories recorded, the most frequently recorded was used, or the most recent, if frequencies were equal. For patients registered in England, if ethnicity was not coded in CPRD, ethnicity data were sourced from linked Hospital Episode Statistics (HES) data,^27^ where available. Geographic region refers to the location of the primary care practice at which the patient was registered at - and included Northern Ireland, Scotland, Wales, and nine regions across England (defined according to Office for National Statistics categories). Linked small area-level data were used for patients registered in England to provide information on relative deprivation (quintile of the 2019 English Index of Multiple Deprivation), derived according to patients’ residential postcode (or the primary care practice postcode as a proxy, if not available). Where a patient had more than one SMI diagnosis recorded over time, the most recent diagnostic category was used as we considered this more likely to be accurate given a more complete clinical history, retaining the first diagnosis date.^28^ We used binary indicators for prescriptions of antidepressants and mood stabilisers (defined according to British National Formulary [BNF] chapters 4.3 and 4.2.3,^20^ respectively) during the study period. Follow-up time was calculated as the amount of time in years that patients were registered in primary care during the study period (accounting for end of registration, death, or administrative censoring).

### Statistical analysis

All analyses were conducted in R (version 4.3.1), with code available in the online repository (https://github.com/Alvin-RB/antipsychotics_descriptive_study_cprd). Descriptive statistics were used to characterise the cohort, stratified by antipsychotic exposure status (objective one). To describe antipsychotic prescribing trends (objective two), we first calculated the number of patients prescribed each antipsychotic at least once - overall and separately for long-acting injectables, and reported data for antipsychotics prescribed to ≥50 patients. We then calculated period prevalence rates for the prescribing of antipsychotics, overall and for each antipsychotic, standardised to 1,000 patients, for each year 2000-2019. Within each year, the numerator was the number of patients that received at least one prescription for the antipsychotic over a denominator of the number of patients alive, diagnosed with SMI, and remaining registered in primary care. Allowing for recording delays, diagnosis could be recorded up to two calendar years after prescription to be considered “diagnosed” in the given year. Period prevalence rates for the top 15 most frequently prescribed antipsychotic medications were represented using line graphs, overall and stratified by SMI diagnosis. Line graphs were also used to visualise the mean total daily prescribed oral antipsychotic dose (with 95% confidence intervals) for up to the first 12 prescription dates, stratified by diagnosis, ethnicity, age, sex, and deprivation (objective three). Among those prescribed an antipsychotic more than once, we focused on the first 12 prescription dates amongst patients identified as new users of antipsychotics in the study period to ensure comparable prescribing periods across patients. Assuming an average prescription duration of 28-30 days, we anticipated that this would approximate patients’ first year of prescribing. If the number of daily doses prescribed was missing for a given prescription, it was imputed (Supplementary Table 2). For missing daily dose values, the previous dose was carried forward for the missing observation, only if the dose at the subsequent time-point was the same.

## RESULTS

### Objective one: Characteristics of patients prescribed and not prescribed antipsychotics in primary care

From a total of 514,526 patients ever receiving a SMI diagnosis in the CPRD database during the study period, 309,378 were identified as having an SMI diagnosis first recorded in their primary care record between 2000-2017. From these, 212,618 (68.7%) were prescribed an antipsychotic in primary care at least once between 2000-2019, whilst 96,760 (31.3%) were not (Table 1).

**Table 1.**
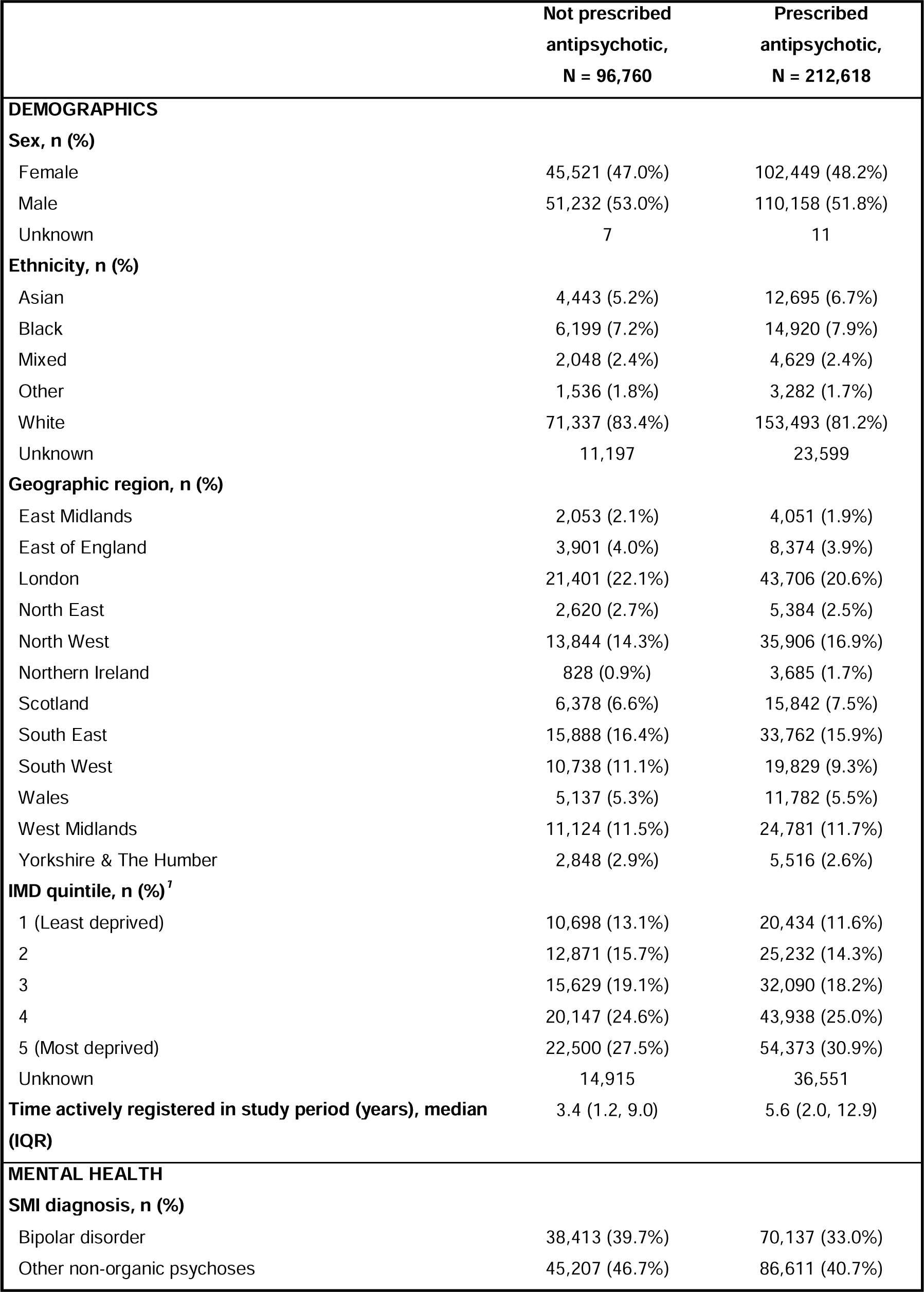
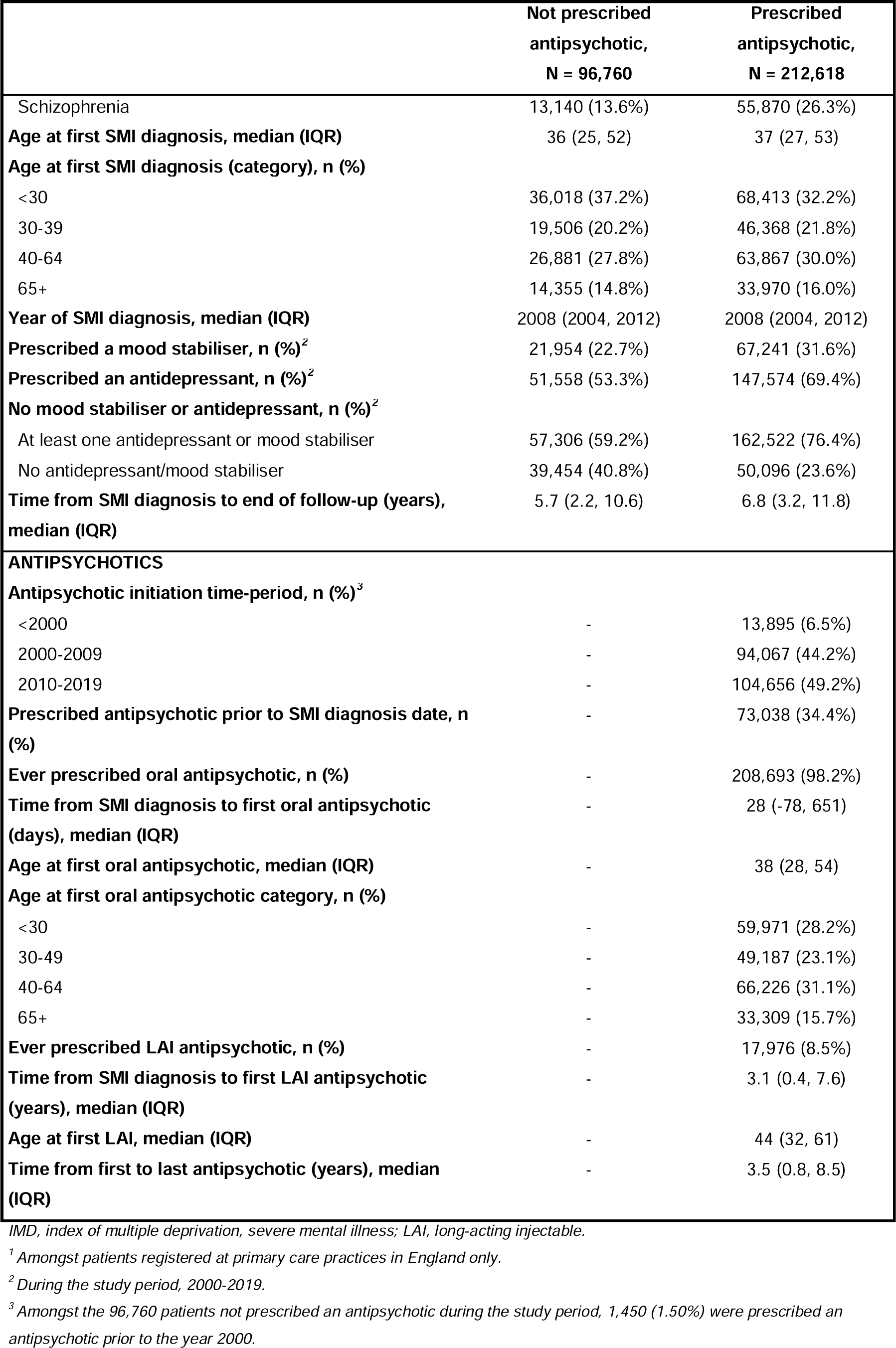
Characteristics of patients prescribed and not prescribed antipsychotics in primary care between 2000-2019.

|  | **Not prescribed**<br>**antipsychotic,**<br>**N = 96,760** | **Prescribed**<br>**antipsychotic,**<br>**N = 212,618** |
| --- | --- | --- |
| **DEMOGRAPHICS** |  |  |
| **Sex, n (%)** |  |  |
| Female | 45,521 (47.0%) | 102,449 (48.2%) |
| Male | 51,232 (53.0%) | 110,158 (51.8%) |
| Unknown | 7 | 11 |
| **Ethnicity, n (%)** |  |  |
| Asian | 4,443 (5.2%) | 12,695 (6.7%) |
| Black | 6,199 (7.2%) | 14,920 (7.9%) |
| Mixed | 2,048 (2.4%) | 4,629 (2.4%) |
| Other | 1,536 (1.8%) | 3,282 (1.7%) |
| White | 71,337 (83.4%) | 153,493 (81.2%) |
| Unknown | 11,197 | 23,599 |
| **Geographic region, n (%)** |  |  |
| East Midlands | 2,053 (2.1%) | 4,051 (1.9%) |
| East of England | 3,901 (4.0%) | 8,374 (3.9%) |
| London | 21,401 (22.1%) | 43,706 (20.6%) |
| North East | 2,620 (2.7%) | 5,384 (2.5%) |
| North West | 13,844 (14.3%) | 35,906 (16.9%) |
| Northern Ireland | 828 (0.9%) | 3,685 (1.7%) |
| Scotland | 6,378 (6.6%) | 15,842 (7.5%) |
| South East | 15,888 (16.4%) | 33,762 (15.9%) |
| South West | 10,738 (11.1%) | 19,829 (9.3%) |
| Wales | 5,137 (5.3%) | 11,782 (5.5%) |
| West Midlands | 11,124 (11.5%) | 24,781 (11.7%) |
| Yorkshire & The Humber | 2,848 (2.9%) | 5,516 (2.6%) |
| **IMD quintile, n (%)<sup>1</sup>** |  |  |
| 1 (Least deprived) | 10,698 (13.1%) | 20,434 (11.6%) |
| 2 | 12,871 (15.7%) | 25,232 (14.3%) |
| 3 | 15,629 (19.1%) | 32,090 (18.2%) |
| 4 | 20,147 (24.6%) | 43,938 (25.0%) |
| 5 (Most deprived) | 22,500 (27.5%) | 54,373 (30.9%) |
| Unknown | 14,915 | 36,551 |
| **Time actively registered in study period (years), median (IQR)** | 3.4 (1.2, 9.0) | 5.6 (2.0, 12.9) |
| **MENTAL HEALTH** |  |  |
| **SMI diagnosis, n (%)** |  |  |
| Bipolar disorder | 38,413 (39.7%) | 70,137 (33.0%) |
| Other non-organic psychoses | 45,207 (46.7%) | 86,611 (40.7%) |
| Schizophrenia | 13,140 (13.6%) | 55,870 (26.3%) |
| **Age at first SMI diagnosis, median (IQR)** | 36 (25, 52) | 37 (27, 53) |
| **Age at first SMI diagnosis (category), n (%)** |  |  |
| <30 | 36,018 (37.2%) | 68,413 (32.2%) |
| 30-39 | 19,506 (20.2%) | 46,368 (21.8%) |
| 40-64 | 26,881 (27.8%) | 63,867 (30.0%) |
| 65+ | 14,355 (14.8%) | 33,970 (16.0%) |
| **Year of SMI diagnosis, median (IQR)** | 2008 (2004, 2012) | 2008 (2004, 2012) |
| **Prescribed a mood stabiliser, n (%)<sup>2</sup>** | 21,954 (22.7%) | 67,241 (31.6%) |
| **Prescribed an antidepressant, n (%)<sup>2</sup>** | 51,558 (53.3%) | 147,574 (69.4%) |
| **No mood stabiliser or antidepressant, n (%)<sup>2</sup>** |  |  |
| At least one antidepressant or mood stabiliser | 57,306 (59.2%) | 162,522 (76.4%) |
| No antidepressant/mood stabiliser | 39,454 (40.8%) | 50,096 (23.6%) |
| **Time from SMI diagnosis to end of follow-up (years), median (IQR)** | 5.7 (2.2, 10.6) | 6.8 (3.2, 11.8) |
| **ANTIPSYCHOTICS** |  |  |
| **Antipsychotic initiation time-period, n (%)<sup>3</sup>** |  |  |
| <2000 | - | 13,895 (6.5%) |
| 2000-2009 | - | 94,067 (44.2%) |
| 2010-2019 | - | 104,656 (49.2%) |
| **Prescribed antipsychotic prior to SMI diagnosis date, n (%)** | - | 73,038 (34.4%) |
| **Ever prescribed oral antipsychotic, n (%)** | - | 208,693 (98.2%) |
| **Time from SMI diagnosis to first oral antipsychotic (days), median (IQR)** | - | 28 (-78, 651) |
| **Age at first oral antipsychotic, median (IQR)** | - | 38 (28, 54) |
| **Age at first oral antipsychotic category, n (%)** |  |  |
| <30 | - | 59,971 (28.2%) |
| 30-49 | - | 49,187 (23.1%) |
| 40-64 | - | 66,226 (31.1%) |
| 65+ | - | 33,309 (15.7%) |
| **Ever prescribed LAI antipsychotic, n (%)** | - | 17,976 (8.5%) |
| **Time from SMI diagnosis to first LAI antipsychotic (years), median (IQR)** | - | 3.1 (0.4, 7.6) |
| **Age at first LAI, median (IQR)** | - | 44 (32, 61) |
| **Time from first to last antipsychotic (years), median (IQR)** | - | 3.5 (0.8, 8.5) |

IMD, index of multiple deprivation, severe mental illness; LAI, long-acting injectable.
<sup>1</sup> Amongst patients registered at primary care practices in England only.
<sup>2</sup> During the study period, 2000-2019.
<sup>3</sup> Amongst the 96,760 patients not prescribed an antipsychotic during the study period, 1,450 (1.50%) were prescribed an antipsychotic prior to the year 2000.

Patients prescribed and not prescribed antipsychotics were broadly similar demographically (Supplementary Figure 2), but some regional differences were observed - with greater proportions prescribed antipsychotics in the North West of England, Northern Ireland, and Wales. The proportion prescribed antipsychotics was higher amongst patients diagnosed with schizophrenia (81.0%) than with bipolar disorder (64.6%) and other non-organic psychoses (65.7%). Amongst those not prescribed antipsychotics, over a fifth (22.7%) were prescribed mood stabilisers and over half (53.3%) antidepressants in the study period, but these proportions were higher amongst those prescribed antipsychotics (31.6% and 69.4%, respectively). The median time registered in primary care during the study period was shorter amongst those not receiving antipsychotics (3.4 vs. 5.6 years). Comparisons are stratified by SMI diagnosis in Supplementary Tables 3-5.

Among those prescribed antipsychotics, almost all (98.2%) received at least one oral prescription. The median time from SMI diagnosis to first oral antipsychotic prescription was 28 (IQR, −78 to 651) days and from first to most recent or last antipsychotic prescription was 3.5 (IQR, 0.8 to 8.5) years. Over a third (34.4%) were prescribed an antipsychotic a median (IQR) of 16 (3 to 53) months prior to having an SMI diagnosis recorded in their primary care record. Of those prescribed an antipsychotic overall, 8.5% were prescribed a long-acting injectable, but this proportion ranged from 4.5% to 16.4% amongst those diagnosed with bipolar disorder and schizophrenia, respectively (Supplementary Tables 3-5). Stratified by ethnicity, the proportion prescribed a long-acting injectable was highest amongst Black patients (9.2%), very similar amongst Asian and Mixed patients (6.8% and 6.9%, respectively) and lowest amongst White patients and those of other ethnicities (5.5% and 4.5%, respectively).

### Objective two: Antipsychotic prescribing trends

After excluding 1,171 prescriptions (across 764 patients) considered potentially erroneous duplicates, the 212,618 patients diagnosed with SMI and prescribed an antipsychotic had a total of 11,745,996 prescriptions, covering 33 different medications, between 2000-2019. Olanzapine was prescribed at least once to 91,961 (43.3%) patients and was the most frequently prescribed, followed by quetiapine (n=70,250, 33.0%), risperidone (n=63,893, 30.1%), and aripiprazole (n=44,344, 20.9%) (Supplementary Figure 3). These four antipsychotics accounted for 78.8% of all prescriptions. The most frequently prescribed first-generation antipsychotics were chlorpromazine (n=17,195, 8.1%) and haloperidol (n=17,119, 8.1%). Clozapine was infrequently prescribed in primary care (n=5,346, 2.5%). Trends were similar when considering first- and second-line medications (Supplementary Table 6). The most frequently prescribed long-acting injectables were flupentixol and zuclopenthixol (Supplementary Figure 4).

The overall prevalence of antipsychotic prescribing was 426 (95% CI, 420 to 433) per 1,000 patients in the year 2000, reaching a peak of 550 (95% CI, 547 to 553) in 2016, then decreasing to 470 (95% CI, 468 to 473) in 2019 (Supplementary Figure 5). Annual prevalence rates for individual antipsychotics varied over time (Supplementary Figure 6) and according to SMI diagnosis. Amongst patients with a diagnosis of schizophrenia, olanzapine was most frequently prescribed, and, for most of the time-period, this was followed by risperidone (Figure 1).

However, in 2015, aripiprazole overtook risperidone. Amongst those with a diagnosis of bipolar disorder, olanzapine had been the most frequently prescribed up until to 2009, after which it was overtaken by quetiapine (Figure 2). Amongst patients diagnosed with other non-organic psychoses, prescribing prevalences for quetiapine, aripiprazole, and risperidone were all relatively similar since 2016, but olanzapine was the most frequently prescribed throughout (Supplementary Figure 7).

**Figure 1.**
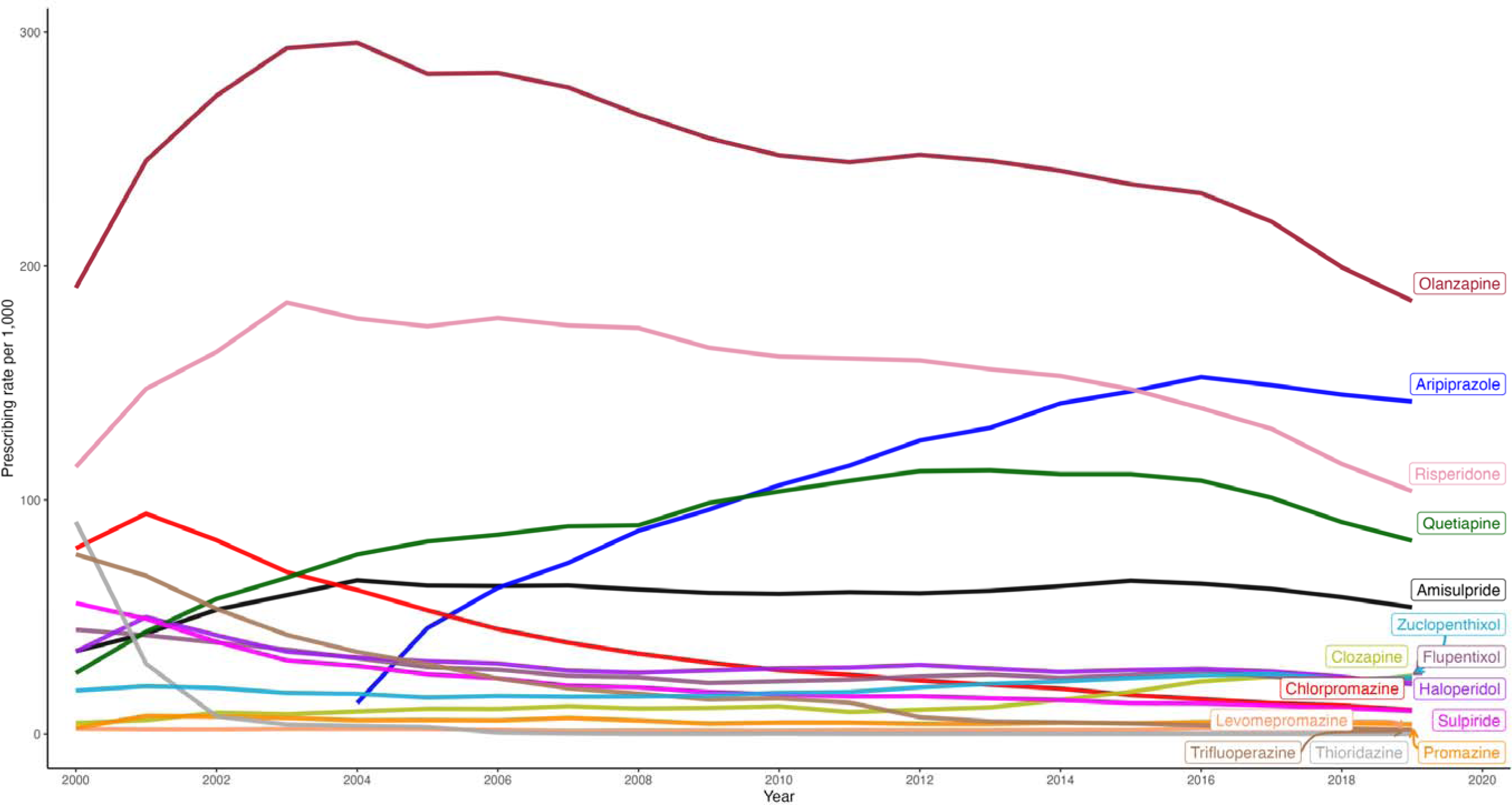
Annual prevalence rates for the prescribing of antipsychotics to patients diagnosed with schizophrenia.

**Figure 2.**
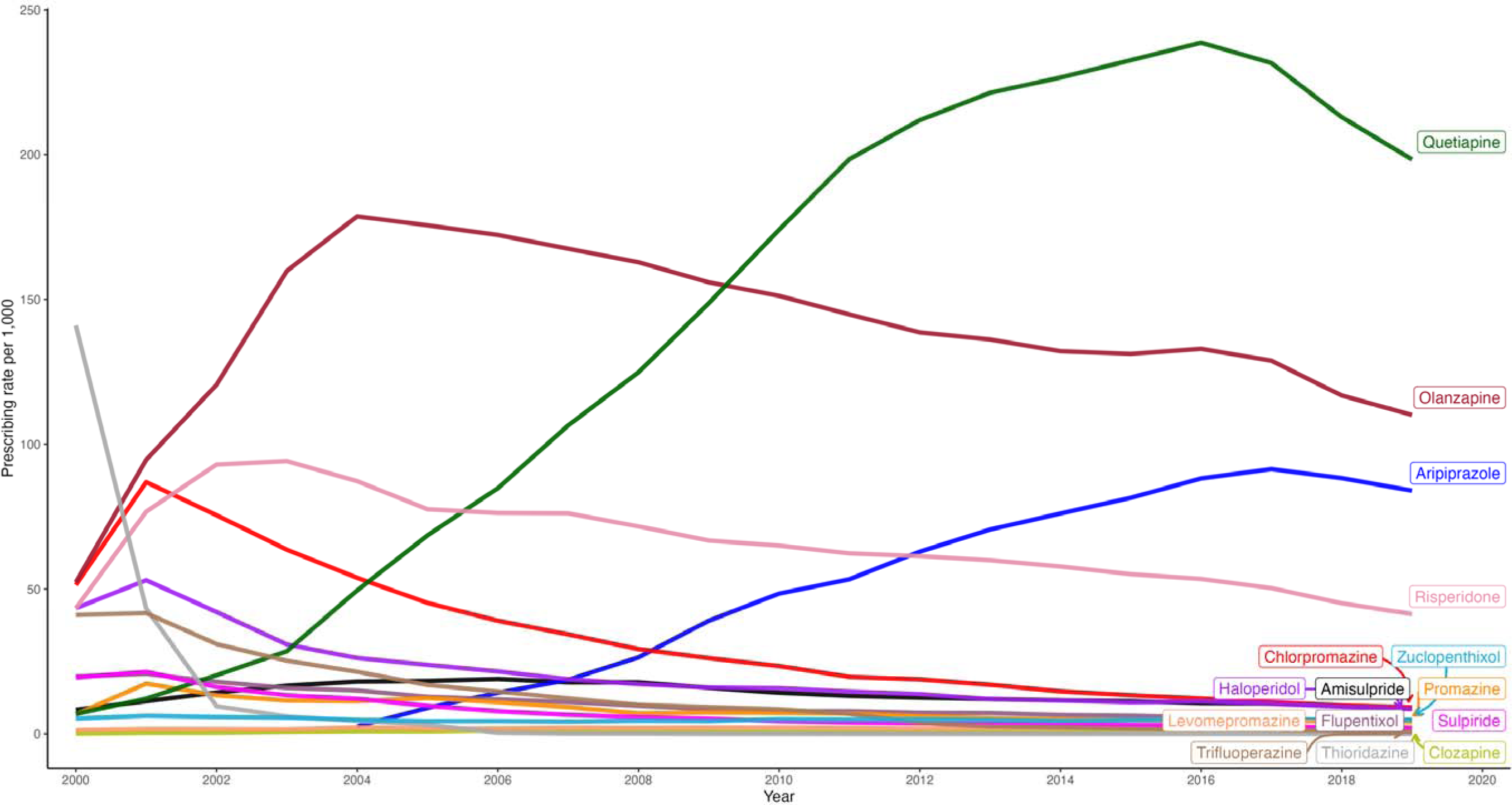
Annual prevalence rates for the prescribing of antipsychotics to patients diagnosed with bipolar disorder.

### Objective three: Variation in average prescribed daily antipsychotic dose over patients’ first year of prescribing

Of the 212,618 patients prescribed antipsychotics between 2000-2019, 194,979 were identified as newly prescribed an oral antipsychotic in primary care during the study period. After exclusions (15,703 for receiving just one prescription and 42 due to having no known doses across their first 12 prescription dates), a total of 179,234 patients, with 1,780,077 prescription dates, were included.

Mean total daily prescribed oral antipsychotic doses varied across subgroups, but all tended to increase slightly over the first 12 prescription dates. Stratified by SMI diagnosis, patients diagnosed with schizophrenia were prescribed the highest doses (mean [SD] daily dose at 12^th^ prescription date: 10.7 [7.4] mg olanzapine equivalent dose), whilst those diagnosed with bipolar disorder were prescribed the lowest doses (7.2 [6.0] mg) (Figure 3). When stratified by ethnicity, Black patients were prescribed the highest doses (9.7 [6.9] mg olanzapine equivalent dose), followed by Mixed (9.5 [6.7] mg), Other (9.0 [6.6] mg), then Asian (8.8 [6.7] mg), whilst White patients were prescribed the lowest doses (8.1 [6.7] mg) [doses were lower in those with missing ethnicity data (7.7 [6.5] mg), but very similar to those of White patients] (Figure 3). Mean daily doses were higher in males compared to females (Supplementary Figure 8), in younger compared to older (65+) patients (Supplementary Figure 9), and in patients in more versus less deprived areas (Supplementary Figure 10).

**Figure 3.**
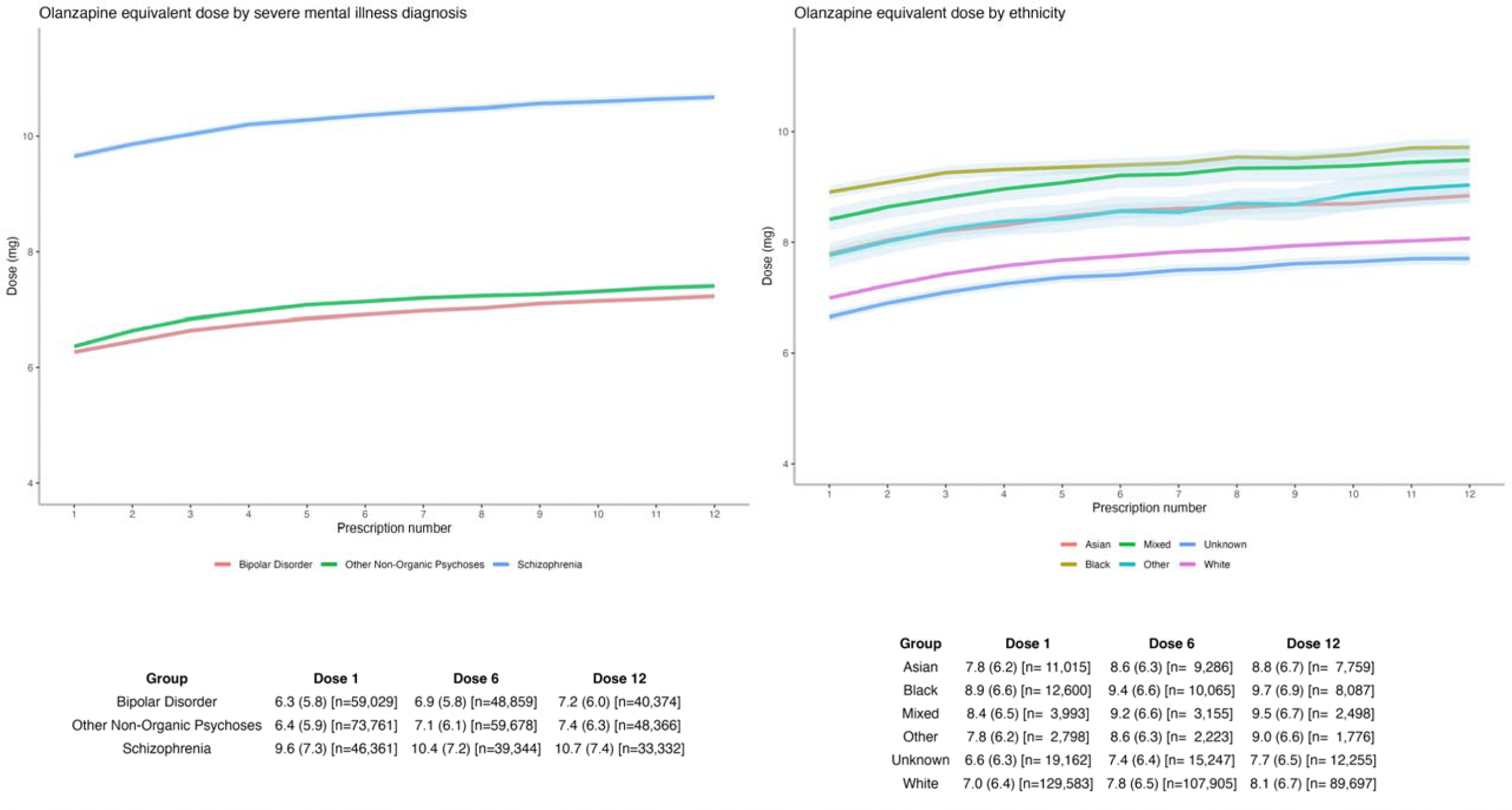
Mean total daily prescribed oral antipsychotic dose over the first 12 prescriptions – stratified by severe mental illness diagnosis and ethnicity.

## DISCUSSION

Using a large, longitudinal sample of 309,378 patients diagnosed with SMI between 2000-2017, we provide contemporary data (2000-2019) on antipsychotic prescribing practice in UK primary care. We identify several important findings relevant to informing future quality improvement and research into safer prescribing, including (1) prescribing is dominated by olanzapine, quetiapine, risperidone, and aripiprazole - accounting for 79% of all prescriptions; (2) disparities in prescribed antipsychotic and dose exist - namely higher doses prescribed to patients with characteristics such as ethnic minority status and greater deprivation and (3) almost a third of patients with a contemporaneous SMI diagnosis are not prescribed antipsychotics in primary care.

Overall, olanzapine was the most frequently prescribed antipsychotic throughout the study period. Stratified by diagnosis, this remained true for schizophrenia and other non-organic psychoses, but not for bipolar disorder, where, since 2010, quetiapine was most frequently prescribed. Adverse cardiometabolic effects are a major concern of second-generation antipsychotics and when antipsychotics are ranked according to their impact on cardiometabolic parameters, olanzapine is consistently identified as one of the worst-ranking, particularly for changes in body weight, body mass index, and low-density lipoprotein cholesterol.^3^ The continued popularity of olanzapine may be due to a perceived greater efficacy compared to other antipsychotics,^29^ despite most antipsychotics being considered broadly comparable in efficacy.^30^ Alternatively, for patients well-established on olanzapine, it may be due to the perceived relapse risk presented by switching to a different antipsychotic with less cardiometabolic burden.

The 2004 licensing of aripiprazole led to a major change in prescribing, whereby prescriptions of aripiprazole have increased year-on-year - now making aripiprazole one of the most frequently prescribed antipsychotics. This is an important development as current evidence suggests that aripiprazole is associated with less adverse cardiometabolic effects, especially when compared to olanzapine and quetiapine.^3,31^ However, some reviews have reported aripiprazole to be less efficacious than some antipsychotics, such as olanzapine and risperidone,^29,32^ although others report no differences.^30^ Aripiprazole is also suggested to exacerbate psychotic symptoms amongst patients with significant prior antipsychotic exposure.^33^ Aripiprazole is still one of the most recently licensed antipsychotics and current popularity might reflect effectiveness of pharmaceutical marketing or a novelty effect in the face of limited innovations in the development of new antipsychotics. These issues highlight the difficulty, but necessity, of evaluating the risk/benefit ratio of individual antipsychotics.

If it were possible to optimise current prescribing, then efforts focusing on olanzapine, quetiapine, risperidone, and aripiprazole could have a large impact on the SMI population given their very widespread use (79% of all antipsychotic prescriptions). Studies of the comparative safety and effectiveness of aripiprazole are particularly warranted given aripiprazole’s potential to reduce cardiometabolic risk alongside concerns of possibly lesser effectiveness. Conversely, some antipsychotics are rarely prescribed and so there is limited opportunity to learn about their relative risks and benefits in pharmacoepidemiologic studies using routine clinical data.

To inform future quality improvement and research, we sought to describe current practice and identify subgroups that may potentially be at more risk of dose-dependent adverse reactions. We found that, on average, higher doses were prescribed to patients with the following characteristics: diagnosis of schizophrenia, ethnic minority status, male sex, younger age, and greater deprivation. In addition, we replicated higher use of long-acting injectables amongst Black patients.^12,34^ To our knowledge, this is the first study to report disparities according to ethnicity and deprivation in prescribed antipsychotic dose in UK primary care. For ethnicity - patients from all ethnic minorities were prescribed higher doses than White patients. Confidence intervals for ethnic minority groups were inevitably wider than, but never overlapped with, the White group - owing to smaller sample sizes reflecting minority status. We did not aim to estimate whether certain characteristics are *causally* related to being prescribed higher doses or to identify potential mediating factors, and therefore our analyses were unadjusted, as recommended for descriptive studies.^35^ Clearly, multiple factors may influence decisions to prescribe at a certain dose, and further research is needed to disentangle the effects of these factors in order to explain, and potentially inform efforts to reduce, these disparities. Causal inference approaches accounting for a wide range of potential confounders (e.g., markers of severity, access to care care), alongside qualitative approaches examining clinical decision-making, would be informative.

Finally, there was a trend of declining antipsychotic prescribing rates over the later study years and, overall, almost one third of patients with a contemporaneous SMI diagnosis were not prescribed antipsychotics in primary care. This is potentially concerning given reports of worse outcomes, including higher mortality, amongst patients diagnosed with schizophrenia-spectrum disorders that are not prescribed antipsychotics.^2^ Noting that less than 2% of these patients were identified as prescribed an antipsychotic in primary care prior to the study period, It is difficult to ascertain if the remainder were truly unexposed based on primary care records alone. Although the shorter follow-up time reduced the opportunity to identify antipsychotic prescriptions, this group still had a median follow-up 3.4 years - seemingly sufficient to detect regular prescribing. Nevertheless, a small proportion will likely have been prescribed antipsychotics exclusively in secondary care (e.g., as inpatients) - an issue particularly relevant for clozapine and long-acting injectables. Alternative explanations might include: patients declining antipsychotics and/or instead receiving psychological interventions or non-antipsychotic pharmacotherapies (particularly for those diagnosed with bipolar disorder); patients with brief or less severe psychotic episodes; or perhaps some were not in contact with services following diagnosis (although many were prescribed other psychiatric medications).

### Strengths and limitations

A major strength of this study is the large longitudinal cohort of patients diagnosed with SMI, derived from CPRD - which is broadly representative of the UK population.^14,15^ CPRD includes all prescriptions issued in primary care and is therefore accurate in terms of planned treatment, and prescriptions issued repeatedly suggest adherence to that medication. We covered a 20-year period, enabling the identification of contemporary trends in prescribing for SMI, whereas other recent studies focused on other diagnoses or on all-cause prescribing. When analysing antipsychotic dose, it was important to consider dose over multiple time-points in order to capture potential changes, rather than just the starting dose, which may not have accurately reflected ongoing management.

This study also has limitations. First, we included only prescriptions issued from primary care (and so were not able to comment in detail on clozapine prescribing) and did not have data on dispensing or individual patient adherence (although repeat prescriptions issued with a regular cadence suggest adherence). Studies combining prescribing and dispensing data across primary and secondary care are needed to characterise the complete national picture on antipsychotic prescribing; such studies might soon be feasible with the continued development of national data resources.^36^ Second, we studied broad ethnic groups, consistent with UK-census high-level ethnicity categories, and focused on between-group, rather than within-group, heterogeneity. Studies of more specific ethnic groups are needed, but will be challenging due to smaller sample sizes and greater misclassification risk. Moreover, although ethnicity should be self-reported in primary care, we cannot verify this assumption. Third, the study period went up to 2019 and therefore did not cover the COVID-19 pandemic period. Initial evidence from an England-wide analysis suggests that antipsychotic prescribing remained relatively stable in the SMI population during the pandemic period,^37^ but studies with a greater SMI focus are warranted.

## Conclusion

Antipsychotic prescribing is dominated by olanzapine, quetiapine, risperidone, and aripiprazole. Two thirds of patients with diagnosed SMI were prescribed antipsychotics in primary care, but this proportion varied according to SMI diagnosis. There were disparities in both receipt and dose of antipsychotics across subgroups - further efforts are needed to understand why certain groups are prescribed higher doses and whether they require dose optimisation to minimise side effects.

## Data Availability

Data underlying this study were accessed via Clinical Practice Research Datalink (CPRD) under approved protocol no. 21_000729. Authors are not able to share the data directly, however data can be accessed directly from Clinical Practice Research Datalink (CPRD) following approval and licensing.

https://cprd.com/

## Acknowledgements

This study is based in part on data from the Clinical Practice Research Datalink obtained under licence from the UK Medicines and Healthcare products Regulatory Agency. The data is provided by patients and collected by the NHS as part of their care and support. The interpretation and conclusions contained in this study are those of the authors alone.

## Data availability

Data underlying this study were accessed via Clinical Practice Research Datalink (CPRD) under approved protocol no. 21_000729. Authors are not able to share the data directly, however data can be accessed directly from Clinical Practice Research Datalink (CPRD) following approval and licensing (see https://cprd.com/ for further details).

## Analytic code availability

The analytic code supporting the findings are available in the online repository (https://github.com/Alvin-RB/antipsychotics_descriptive_study_cprd).

## Author contributions

ARB, EB, DPJO, and JFH forumated the research questions and designed the study. ARB analysed the data. ARB wrote the first draft of the manuscript and NL, SH, KKCM, EB, DPJO and JFH critically reviewed the manuscript for important intellectual content. All authors approved the final version to be published and agree to be accountable for all aspects of the work in ensuring that questions related to the accuracy or integrity of any part of the work are appropriately investigated and resolved.

