## Supplementary Material for "Prescribing of antipsychotics for people diagnosed with severe mental illness in UK primary care: A 20-year investigation of who receives treatment, with which agents, and at what doses"

### Supplementary Tables

#### Supplementary Table 1. List of antipsychotics.

| Generic name | In BNF | In WHO ATC | Brand names and Alternative spellings |
|---|---|---|---|
| Acepromazine | | ✓ | Acetopromazine, Acetylpromazine |
| Acetophenazine | | ✓ | Tindal |
| Amisulpride | ✓ | ✓ | Solian, Socian, Deniban |
| Aripiprazole | ✓ | ✓ | Abilify, Aristada, Arpoya |
| Asenapine | ✓ | ✓ | Sycrest, Saphris, Secuado |
| Benperidol | ✓ | ✓ | Anquil, Frenactil |
| Blonanserin | | | Lonasen |
| Brexpiprazole | | ✓ | Rexulti, Rxulti |
| Bromperidol | | ✓ | Bromidol, Bromodol, Erodium, Impromen, Tesoprel |
| Butaperazine | | ✓ | Repoise, Tyrylen |
| Cariprazine | ✓ | | Vraylar, Reagila |
| Carpipramine | | | Prazinil, Defekton |
| Chlorproethazine | | ✓ | Neuriplege |
| Chlorpromazine | ✓ | ✓ | Largactil, Thorazine |
| Chlorprothixene | ✓ | ✓ | Truxal |
| Clocapramine | | | |
| Clopenthixol | | ✓ | Clopentixol, Sordinol |
| Clotiapine | | ✓ | Etumina, Etumine, Entumin, Etomine, Entumine |
| Clozapine | ✓ | ✓ | Clozaril, FazaClo, Denzapine, Leponex, Versacloz, Zaponex |
| Cyamemazine | | ✓ | Cyamepromazine, Tercian |
| Dixyrazine | | ✓ | Dixypazin, Ansiolene, Esocalm, Esucos, Metronal, Roscal |

| | | | |
|---|---|---|---|
| Droperidol | | ✓ | Inapsine, Droleptan, Dridol, Xomolix |
| Fluanisone | | ✓ | |
| Flupentixol | ✓ | ✓ | Depixol, Fluanxol, Flupenthixol, Psytixol |
| Fluphenazine | ✓ | ✓ | Modecate, Prolixin, Moditen |
| Fluspirilene | | ✓ | Imap, Redeptin |
| Haloperidol | ✓ | ✓ | Haldol, Serenace, Dozic, Halkid |
| Iloperidone | | ✓ | Fanapt, Zomaril |
| Levomepromazine | ✓ | ✓ | Nozinan, Levoprome, Detenler, Hirnamin, Levotomin, Neurocil, Methotrimeprazine |
| Levosulpiride | | ✓ | Neoprad |
| Loxapine | ✓ | ✓ | Loxitane, Adasuve |
| Lumateperone | | ✓ | Caplyta |
| Lurasidone | | ✓ | Latuda |
| Melperone | ✓ | ✓ | Buronil, Bunil, Eunerpan |
| Mesoridazine | | ✓ | Serentil |
| Molindone | | ✓ | Moban |
| Moperone | | ✓ | Luvatren |
| Mosapramine | | ✓ | Cremin |
| Olanzapine | ✓ | ✓ | Zyprexa, Zalasta, Zypadhera, Arkolamyl |
| Oxypertine | | ✓ | Equipertine, Forit, Integrin, Lanturil, Lotawin, Opertil |
| Paliperidone | ✓ | ✓ | Invega, Xeplion, Trevicta |
| Penfluridol | | ✓ | Semap, Micefal, Longoperidol |
| Perazine | | ✓ | Taxilan |
| Pericyazine | ✓ | ✓ | Neulactil, Periciazine, Propericiazine |
| Perospirone | | | Lullan |
| Perphenazine | ✓ | ✓ | Fentazin, Trilafon |

| | | | |
|---|---|---|---|
| Pimavanserin | | ✓ | Nuplazid |
| Pimozide | ✓ | ✓ | Orap |
| Pipamperone | | ✓ | Dipiperon, Dipiperal, Piperonil, Piperonyl, Propitan, Carpiperone, Floropipamide, Fluoropipamide, Floropipamide |
| Pipotiazine | | ✓ | Piportil, Pipothiazine |
| Promazine | ✓ | ✓ | Sparine |
| Prothipendyl | | ✓ | Dominal, Timovan, Tolnate, Azapromazine, Phrenotropin |
| Quetiapine | ✓ | ✓ | Seroquel, Temprolide, Tenprolide, Atrolak, Biquelle, Brancico, Sondate, Zaluron, Mintreleq, Psyquet |
| Remoxipride | | ✓ | Roxiam |
| Risperidone | ✓ | ✓ | Risperdal, Okedi |
| Sertindole | | ✓ | Serdolect, Serlect |
| Sulpiride | ✓ | ✓ | Dolmatil, Dogmatil, Sulpor |
| Sultopride | | ✓ | Barnetil, Barnotil, Topral |
| Thiopropazate | | ✓ | Artalan, Dartal, Dartalan, Dartan |
| Thioproperazine | | ✓ | Majeptil |
| Thioridazine | ✓ | ✓ | Melleril, Mellaril |
| Tiapride | | ✓ | Tiapridal |
| Tiotixene | | ✓ | Thiothixene, Navane |
| Trifluoperazine | ✓ | ✓ | Stelazine, Eskazinyl, Eskazine, Jatroneural, Terrazine |
| Trifluperidol | | ✓ | Triperidol |
| Triflupromazine | | ✓ | |
| Veralipride | | ✓ | Agreal, Agradil |
| Ziprasidone | | ✓ | Zeldox, Geodon, Zipwell |
| Zotepine | ✓ | | Nipolept, Losizopilon, Zoleptil, Zotepin |
| Zuclopenthixol | ✓ | ✓ | Clopixol, Cisordinol, Zuclopentixol |

*BNF, British National Formulary; WHO ATC, World Health Organisation Anatomical Therapeutic Chemical Classification System.*

**Supplementary Table 2. Imputation scheme for missing numbers of daily doses.**

| Imputation order | Fields used for imputation |
|---|---|
| 1 | Patient ID, product code |
| 2 | Patient ID, product name |
| 3 | Patient ID, antipsychotic, strength |
| 4 | Patient ID, antipsychotic, numeric dose |
| 5 | Practice ID, SMI diagnosis, product code |
| 6 | Practice ID, SMI diagnosis, product name |
| 7 | Practice ID, SMI diagnosis, antipsychotic, strength |
| 8 | Practice ID, SMI diagnosis, antipsychotic, numeric dose |
| 9 | SMI diagnosis, product code |
| 10 | SMI diagnosis, product name |
| 11 | SMI diagnosis, antipsychotic, strength |
| 12 | SMI diagnosis, antipsychotic, numeric dose |
| 13 | Antipsychotic, strength |
| 14 | Antipsychotic, numeric dose |

*Single imputation was carried out using the impute_ndd function in the drugprepR R package.[1] The imputation scheme started more specific and, if values were still missing, then they were considered for imputation at the next less specific level of the scheme. In total, 299,577 of 1,780,720 (16.8%) number of daily doses (NDDs) were imputed. The imputed NDDs were across 57,163 patients. The median (IQR) of the imputed NDDs was 1 (1-2) which compared to a median IQR of the non-imputed NDDs of 1 (1-2).*

**Supplementary Table 3. Characteristics of patients diagnosed with schizophrenia prescribed and not prescribed antipsychotics in primary care between 2000-2019.**

| | Not prescribed antipsychotic, N = 13,140 | Prescribed antipsychotic, N = 55,870 |
|---|---|---|
| **DEMOGRAPHICS** | | |
| **Sex, n (%)** | | |
| Female | 3,775 (28.7%) | 18,814 (33.7%) |
| Male | 9,362 (71.3%) | 37,055 (66.3%) |
| Unknown | 3 | 1 |
| **Ethnicity, n (%)** | | |
| Asian | 856 (7.4%) | 4,911 (9.7%) |
| Black | 1,946 (16.8%) | 7,040 (13.9%) |
| Mixed | 435 (3.8%) | 1,743 (3.5%) |
| Other | 251 (2.2%) | 991 (2.0%) |
| White | 8,061 (69.8%) | 35,833 (70.9%) |
| Unknown | 1,591 | 5,352 |
| **Geographic region, n (%)** | | |
| East Midlands | 190 (1.4%) | 827 (1.5%) |
| East of England | 431 (3.3%) | 1,792 (3.2%) |
| London | 4,272 (32.5%) | 16,218 (29.0%) |
| North East | 232 (1.8%) | 1,259 (2.3%) |
| North West | 1,805 (13.7%) | 9,981 (17.9%) |
| Northern Ireland | 69 (0.5%) | 815 (1.5%) |
| Scotland | 738 (5.6%) | 3,460 (6.2%) |
| South East | 1,484 (11.3%) | 6,878 (12.3%) |
| South West | 1,142 (8.7%) | 4,215 (7.5%) |
| Wales | 852 (6.5%) | 2,672 (4.8%) |
| West Midlands | 1,648 (12.5%) | 6,444 (11.5%) |
| Yorkshire & The Humber | 277 (2.1%) | 1,309 (2.3%) |
| **IMD quintile, n (%)[1]** | | |
| 1 (Least deprived) | 683 (6.2%) | 3,363 (7.1%) |
| 2 | 1,193 (10.8%) | 5,173 (10.8%) |
| 3 | 1,983 (17.9%) | 8,063 (16.9%) |
| 4 | 3,250 (29.4%) | 13,406 (28.1%) |
| 5 (Most deprived) | 3,962 (35.8%) | 17,689 (37.1%) |
| Unknown | 2,069 | 8,176 |
| **Time actively registered in study period (years), median (IQR)** | 2.1 (0.9, 5.0) | 4.5 (1.7, 10.6) |
| **MENTAL HEALTH** | | |
| **Age at first SMI diagnosis, median (IQR)** | 32 (23, 44) | 34 (25, 46) |
| **Age at first SMI diagnosis (category), n (%)** | | |
| <30 | 5,868 (44.7%) | 21,966 (39.3%) |

| | Not prescribed antipsychotic, N = 13,140 | Prescribed antipsychotic, N = 55,870 |
|---|---|---|
| 30-39 | 3,016 (23.0%) | 13,642 (24.4%) |
| 40-64 | 3,331 (25.4%) | 15,601 (27.9%) |
| 65+ | 925 (7.0%) | 4,661 (8.3%) |
| **Year of SMI diagnosis, median (IQR)** | 2006 (2003, 2010) | 2006 (2003, 2011) |
| **Prescribed a mood stabiliser, n (%)[2]** | 768 (5.8%) | 8,402 (15.0%) |
| **Prescribed an antidepressant, n (%)[2]** | 3,352 (25.5%) | 31,220 (55.9%) |
| **No mood stabiliser or antidepressant, n (%)[2]** | | |
| At least one antidepressant or mood stabiliser | 3,780 (28.8%) | 34,540 (61.8%) |
| No antidepressant/mood stabiliser | 9,360 (71.2%) | 21,330 (38.2%) |
| **Time from SMI diagnosis to end of follow-up (years), median (IQR)** | 5.7 (2.2, 10.7) | 8.0 (3.8, 13.5) |
| **ANTIPSYCHOTICS** | | |
| **Antipsychotic initiation time-period, n (%)** | | |
| <2000 | - | 3,843 (6.9%) |
| 2000-2009 | - | 26,599 (47.6%) |
| 2010-2019 | - | 25,428 (45.5%) |
| **Prescribed antipsychotic prior to SMI diagnosis date, n (%)** | - | 17,046 (30.5%) |
| **Ever prescribed oral antipsychotic, n (%)** | - | 53,713 (96.1%) |
| **Time from SMI diagnosis to first oral antipsychotic (days), median (IQR)** | - | 64 (-34, 1,185) |
| **Age at first oral antipsychotic, median (IQR)** | - | 35 (27, 47) |
| **Age at first oral antipsychotic category, n (%)** | | |
| <30 | - | 17,918 (32.1%) |
| 30-49 | - | 14,797 (26.5%) |
| 40-64 | - | 16,331 (29.2%) |
| 65+ | - | 4,667 (8.4%) |
| **Ever prescribed LAI antipsychotic, n (%)** | - | 9,186 (16.4%) |
| **Time from SMI diagnosis to first LAI antipsychotic (years), median (IQR)** | - | 2.9 (0.2, 7.8) |
| **Age at first LAI, median (IQR)** | - | 40 (31, 53) |
| **Time from first to last antipsychotic (years), median (IQR)** | - | 4.2 (1.1, 9.5) |

*IMD, index of multiple deprivation, severe mental illness; LAI, long-acting injectable.*

*[1] Amongst patients registered at primary care practices in England only.*

*[2] During the study period, 2000-2019.*

**Supplementary Table 4. Characteristics of patients diagnosed with bipolar disorder prescribed and not prescribed antipsychotics in primary care between 2000-2019.**

| | Not prescribed antipsychotic, N = 38,413 | Prescribed antipsychotic, N = 70,137 |
|---|---|---|
| **DEMOGRAPHICS** | | |
| **Sex, n (%)** | | |
| Female | 21,942 (57.1%) | 42,593 (60.7%) |
| Male | 16,471 (42.9%) | 27,540 (39.3%) |
| Unknown | 0 | 4 |
| **Ethnicity, n (%)** | | |
| Asian | 1,382 (4.1%) | 2,778 (4.4%) |
| Black | 1,007 (3.0%) | 2,150 (3.4%) |
| Mixed | 584 (1.7%) | 1,142 (1.8%) |
| Other | 547 (1.6%) | 829 (1.3%) |
| White | 30,533 (89.7%) | 55,719 (89.0%) |
| Unknown | 4,360 | 7,519 |
| **Geographic region, n (%)** | | |
| East Midlands | 882 (2.3%) | 1,500 (2.1%) |
| East of England | 1,832 (4.8%) | 3,181 (4.5%) |
| London | 7,820 (20.4%) | 11,179 (15.9%) |
| North East | 1,102 (2.9%) | 1,825 (2.6%) |
| North West | 5,074 (13.2%) | 11,291 (16.1%) |
| Northern Ireland | 284 (0.7%) | 1,261 (1.8%) |
| Scotland | 2,684 (7.0%) | 5,673 (8.1%) |
| South East | 7,455 (19.4%) | 13,522 (19.3%) |
| South West | 4,242 (11.0%) | 6,488 (9.3%) |
| Wales | 1,703 (4.4%) | 3,935 (5.6%) |
| West Midlands | 4,174 (10.9%) | 8,426 (12.0%) |
| Yorkshire & The Humber | 1,161 (3.0%) | 1,856 (2.6%) |
| **IMD quintile, n (%)[1]** | | |
| 1 (Least deprived) | 5,504 (16.8%) | 8,817 (15.3%) |
| 2 | 6,047 (18.4%) | 9,967 (17.3%) |
| 3 | 6,709 (20.5%) | 11,299 (19.6%) |
| 4 | 7,543 (23.0%) | 13,118 (22.8%) |
| 5 (Most deprived) | 6,974 (21.3%) | 14,405 (25.0%) |
| Unknown | 5,636 | 12,531 |
| **Time actively registered in study period (years), median (IQR)** | 3.9 (1.4, 10.2) | 6.5 (2.3, 14.4) |
| **MENTAL HEALTH** | | |
| **Age at first SMI diagnosis, median (IQR)** | 39 (27, 54) | 40 (28, 53) |
| **Age at first SMI diagnosis (category), n (%)** | | |
| <30 | 11,837 (30.8%) | 19,233 (27.4%) |

| | Not prescribed antipsychotic, N = 38,413 | Prescribed antipsychotic, N = 70,137 |
|---|---|---|
| 30-39 | 7,992 (20.8%) | 15,751 (22.5%) |
| 40-64 | 12,900 (33.6%) | 25,571 (36.5%) |
| 65+ | 5,684 (14.8%) | 9,582 (13.7%) |
| **Year of SMI diagnosis, median (IQR)** | 2008 (2004, 2012) | 2008 (2004, 2013) |
| **Prescribed a mood stabiliser, n (%)[2]** | 18,860 (49.1%) | 47,058 (67.1%) |
| **Prescribed an antidepressant, n (%)[2]** | 24,765 (64.5%) | 55,643 (79.3%) |
| **No mood stabiliser or antidepressant, n (%)[2]** | | |
| At least one antidepressant or mood stabiliser | 29,260 (76.2%) | 64,137 (91.4%) |
| No antidepressant/mood stabiliser | 9,153 (23.8%) | 6,000 (8.6%) |
| **Time from SMI diagnosis to end of follow-up (years), median (IQR)** | 6.1 (2.5, 11.0) | 7.4 (3.8, 12.3) |
| **ANTIPSYCHOTICS** | | |
| **Antipsychotic initiation time-period, n (%)** | | |
| <2000 | - | 5,619 (8.0%) |
| 2000-2009 | - | 28,880 (41.2%) |
| 2010-2019 | - | 35,638 (50.8%) |
| **Prescribed antipsychotic prior to SMI diagnosis date, n (%)** | - | 27,644 (39.4%) |
| **Ever prescribed oral antipsychotic, n (%)** | - | 69,516 (99.1%) |
| **Time from SMI diagnosis to first oral antipsychotic (days), median (IQR)** | - | 21 (-185, 706) |
| **Age at first oral antipsychotic, median (IQR)** | - | 40 (30, 54) |
| **Age at first oral antipsychotic category, n (%)** | | |
| <30 | - | 17,094 (24.4%) |
| 30-49 | - | 16,451 (23.5%) |
| 40-64 | - | 26,494 (37.8%) |
| 65+ | - | 9,477 (13.5%) |
| **Ever prescribed LAI antipsychotic, n (%)** | - | 3,144 (4.5%) |
| **Time from SMI diagnosis to first LAI antipsychotic (years), median (IQR)** | - | 3.8 (0.6, 8.5) |
| **Age at first LAI, median (IQR)** | - | 49 (35, 68) |
| **Time from first to last antipsychotic (years), median (IQR)** | - | 4.0 (0.9, 9.2) |

*IMD, index of multiple deprivation, severe mental illness; LAI, long-acting injectable.*

*[1] Amongst patients registered at primary care practices in England only.*

*[2] During the study period, 2000-2019.*

**Supplementary Table 5. Characteristics of patients diagnosed with other non-organic psychoses prescribed and not prescribed antipsychotics in primary care between 2000-2019.***

| | Not prescribed antipsychotic, N = 45,207 | Prescribed antipsychotic, N = 86,611 |
|---|---|---|
| **DEMOGRAPHICS** | | |
| **Sex, n (%)** | | |
| Female | 19,804 (43.8%) | 41,042 (47.4%) |
| Male | 25,399 (56.2%) | 45,563 (52.6%) |
| Unknown | 4 | 6 |
| **Ethnicity, n (%)** | | |
| Asian | 2,205 (5.5%) | 5,006 (6.6%) |
| Black | 3,246 (8.1%) | 5,730 (7.6%) |
| Mixed | 1,029 (2.6%) | 1,744 (2.3%) |
| Other | 738 (1.8%) | 1,462 (1.9%) |
| White | 32,743 (81.9%) | 61,941 (81.6%) |
| Unknown | 5,246 | 10,728 |
| **Geographic region, n (%)** | | |
| East Midlands | 981 (2.2%) | 1,724 (2.0%) |
| East of England | 1,638 (3.6%) | 3,401 (3.9%) |
| London | 9,309 (20.6%) | 16,309 (18.8%) |
| North East | 1,286 (2.8%) | 2,300 (2.7%) |
| North West | 6,965 (15.4%) | 14,634 (16.9%) |
| Northern Ireland | 475 (1.1%) | 1,609 (1.9%) |
| Scotland | 2,956 (6.5%) | 6,709 (7.7%) |
| South East | 6,949 (15.4%) | 13,362 (15.4%) |
| South West | 5,354 (11.8%) | 9,126 (10.5%) |
| Wales | 2,582 (5.7%) | 5,175 (6.0%) |
| West Midlands | 5,302 (11.7%) | 9,911 (11.4%) |
| Yorkshire & The Humber | 1,410 (3.1%) | 2,351 (2.7%) |
| **IMD quintile, n (%)[1]** | | |
| 1 (Least deprived) | 4,511 (11.9%) | 8,254 (11.7%) |
| 2 | 5,631 (14.8%) | 10,092 (14.3%) |
| 3 | 6,937 (18.3%) | 12,728 (18.0%) |
| 4 | 9,354 (24.6%) | 17,414 (24.6%) |
| 5 (Most deprived) | 11,564 (30.4%) | 22,279 (31.5%) |
| Unknown | 7,210 | 15,844 |
| **Time actively registered in study period (years), median (IQR)** | 3.6 (1.3, 9.5) | 5.7 (2.0, 13.0) |
| **MENTAL HEALTH** | | |
| **Age at first SMI diagnosis, median (IQR)** | 34 (23, 53) | 39 (27, 61) |
| **Age at first SMI diagnosis (category), n (%)** | | |

| | Not prescribed antipsychotic, N = 45,207 | Prescribed antipsychotic, N = 86,611 |
|---|---|---|
| <30 | 18,313 (40.5%) | 27,214 (31.4%) |
| 30-39 | 8,498 (18.8%) | 16,975 (19.6%) |
| 40-64 | 10,650 (23.6%) | 22,695 (26.2%) |
| 65+ | 7,746 (17.1%) | 19,727 (22.8%) |
| **Year of SMI diagnosis, median (IQR)** | 2008 (2004, 2013) | 2009 (2004, 2013) |
| **Prescribed a mood stabiliser, n (%)**[2] | 2,326 (5.1%) | 11,781 (13.6%) |
| **Prescribed an antidepressant, n (%)**[2] | 23,441 (51.9%) | 60,711 (70.1%) |
| **No mood stabiliser or antidepressant, n (%)**[2] | | |
| At least one antidepressant or mood stabiliser | 24,266 (53.7%) | 63,845 (73.7%) |
| No antidepressant/mood stabiliser | 20,941 (46.3%) | 22,766 (26.3%) |
| **Time from SMI diagnosis to end of follow-up (years), median (IQR)** | 5.4 (2.0, 10.3) | 5.7 (2.5, 10.3) |
| **ANTIPSYCHOTICS** | | |
| **Antipsychotic initiation time-period, n (%)** | | |
| <2000 | - | 4,433 (5.1%) |
| 2000-2009 | - | 38,588 (44.6%) |
| 2010-2019 | - | 43,590 (50.3%) |
| **Prescribed antipsychotic prior to SMI diagnosis date, n (%)** | - | 28,348 (32.7%) |
| **Ever prescribed oral antipsychotic, n (%)** | - | 85,464 (98.7%) |
| **Time from SMI diagnosis to first oral antipsychotic (days), median (IQR)** | - | 21 (-49, 359) |
| **Age at first oral antipsychotic, median (IQR)** | - | 39 (28, 60) |
| **Age at first oral antipsychotic category, n (%)** | | |
| <30 | - | 24,959 (28.8%) |
| 30-49 | - | 17,939 (20.7%) |
| 40-64 | - | 23,401 (27.0%) |
| 65+ | - | 19,165 (22.1%) |
| **Ever prescribed LAI antipsychotic, n (%)** | - | 5,646 (6.5%) |
| **Time from SMI diagnosis to first LAI antipsychotic (years), median (IQR)** | - | 2.9 (0.6, 6.8) |
| **Age at first LAI, median (IQR)** | - | 49 (34, 76) |
| **Time from first to last antipsychotic (years), median (IQR)** | - | 2.8 (0.6, 7.2) |

*IMD, index of multiple deprivation, severe mental illness; LAI, long-acting injectable.*

** Other non-organic psychoses included diagnoses such as psychotic episodes, schizoaffective disorders, delusional disorder, and non-organic psychosis not otherwise specified.*

*[1] Amongst patients registered at primary care practices in England only.*

*[2] During the study period, 2000-2019.*

**Supplementary Table 6. Number and percentage of patients receiving a prescription for an antipsychotic as first- and second-line use, overall and by 10-year time periods.***

| Antipsychotic | First-line[1] Overall N = 204,093 | 2000-2009 N = 96,742 | 2010-2019 N = 107,351 | Second-line[2] Overall N = 83,993 | 2000-2009 N = 35,465 | 2010-2019 N = 48,528 |
|---|---|---|---|---|---|---|
| Amisulpride | 7,420 (3.6%) | 3,970 (4.1%) | 3,450 (3.2%) | 4,271 (5.1%) | 2,349 (6.6%) | 1,922 (4.0%) |
| Aripiprazole | 16,947 (8.3%) | 2,968 (3.1%) | 13,979 (13.0%) | 15,541 (18.5%) | 3,349 (9.4%) | 12,192 (25.1%) |
| Chlorpromazine | 8,243 (4.0%) | 6,486 (6.7%) | 1,757 (1.6%) | 3,248 (3.9%) | 2,292 (6.5%) | 956 (2.0%) |
| Clozapine | 2,181 (1.1%) | 623 (0.6%) | 1,558 (1.5%) | 1,192 (1.4%) | 258 (0.7%) | 934 (1.9%) |
| Droperidol | 112 (0.1%) | 112 (0.1%) | - | 26 (0.0%) | 26 (0.1%) | - |
| Flupentixol | 4,170 (2.0%) | 2,584 (2.7%) | 1,586 (1.5%) | 1,907 (2.3%) | 869 (2.5%) | 1,038 (2.1%) |
| Fluphenazine | 546 (0.3%) | 395 (0.4%) | 151 (0.1%) | 215 (0.3%) | 126 (0.4%) | 89 (0.2%) |
| Haloperidol | 7,712 (3.8%) | 4,626 (4.8%) | 3,086 (2.9%) | 3,957 (4.7%) | 1,877 (5.3%) | 2,080 (4.3%) |
| Levomepromazine | 819 (0.4%) | 199 (0.2%) | 620 (0.6%) | 1,026 (1.2%) | 146 (0.4%) | 880 (1.8%) |
| Lurasidone | 113 (0.1%) | - | 113 (0.1%) | 187 (0.2%) | - | 187 (0.4%) |
| Olanzapine | 62,630 (30.7%) | 32,656 (33.7%) | 29,974 (27.9%) | 16,137 (19.2%) | 7,993 (22.5%) | 8,144 (16.8%) |
| Paliperidone | 479 (0.2%) | 8 (0.0%) | 471 (0.4%) | 656 (0.8%) | 11 (0.0%) | 645 (1.3%) |
| Pericyazine | 446 (0.2%) | 307 (0.3%) | 139 (0.1%) | 208 (0.2%) | 130 (0.4%) | 78 (0.2%) |
| Perphenazine | 163 (0.1%) | 143 (0.1%) | 20 (0.0%) | 68 (0.1%) | 62 (0.2%) | 6 (0.0%) |
| Pimozide | 102 (0.0%) | 86 (0.1%) | 16 (0.0%) | 24 (0.0%) | 17 (0.0%) | 7 (0.0%) |
| Pipotiazine | 266 (0.1%) | 139 (0.1%) | 127 (0.1%) | 166 (0.2%) | 68 (0.2%) | 98 (0.2%) |
| Promazine | 2,207 (1.1%) | 1,308 (1.4%) | 899 (0.8%) | 1,370 (1.6%) | 716 (2.0%) | 654 (1.3%) |
| Quetiapine | 40,087 (19.6%) | 11,250 (11.6%) | 28,837 (26.8%) | 16,637 (19.8%) | 6,317 (17.8%) | 10,320 (21.3%) |
| Risperidone | 38,358 (18.8%) | 20,260 (20.9%) | 18,098 (16.8%) | 13,480 (16.0%) | 6,689 (18.9%) | 6,791 (14.0%) |
| Sulpiride | 2,674 (1.3%) | 1,992 (2.1%) | 682 (0.6%) | 886 (1.1%) | 609 (1.7%) | 277 (0.6%) |
| Thioridazine | 1,891 (0.9%) | 1,891 (2.0%) | - | 178 (0.2%) | 178 (0.5%) | - |
| Trifluoperazine | 4,343 (2.1%) | 3,844 (4.0%) | 499 (0.5%) | 1,176 (1.4%) | 972 (2.7%) | 204 (0.4%) |

| | | | | | | |
|---|---|---|---|---|---|---|
| Zuclopenthixol | 2,214 (1.1%) | 896 (0.9%) | 1,318 (1.2%) | 1,338 (1.6%) | 365 (1.0%) | 973 (2.0%) |

** In patients first prescribed antipsychotics in primary care from the year 2000 to 2019.*

[1] *First recorded antipsychotic medication prescribed in primary care.*

[2] *Second recorded antipsychotic medication prescribed in primary care.*

### Supplementary Figures

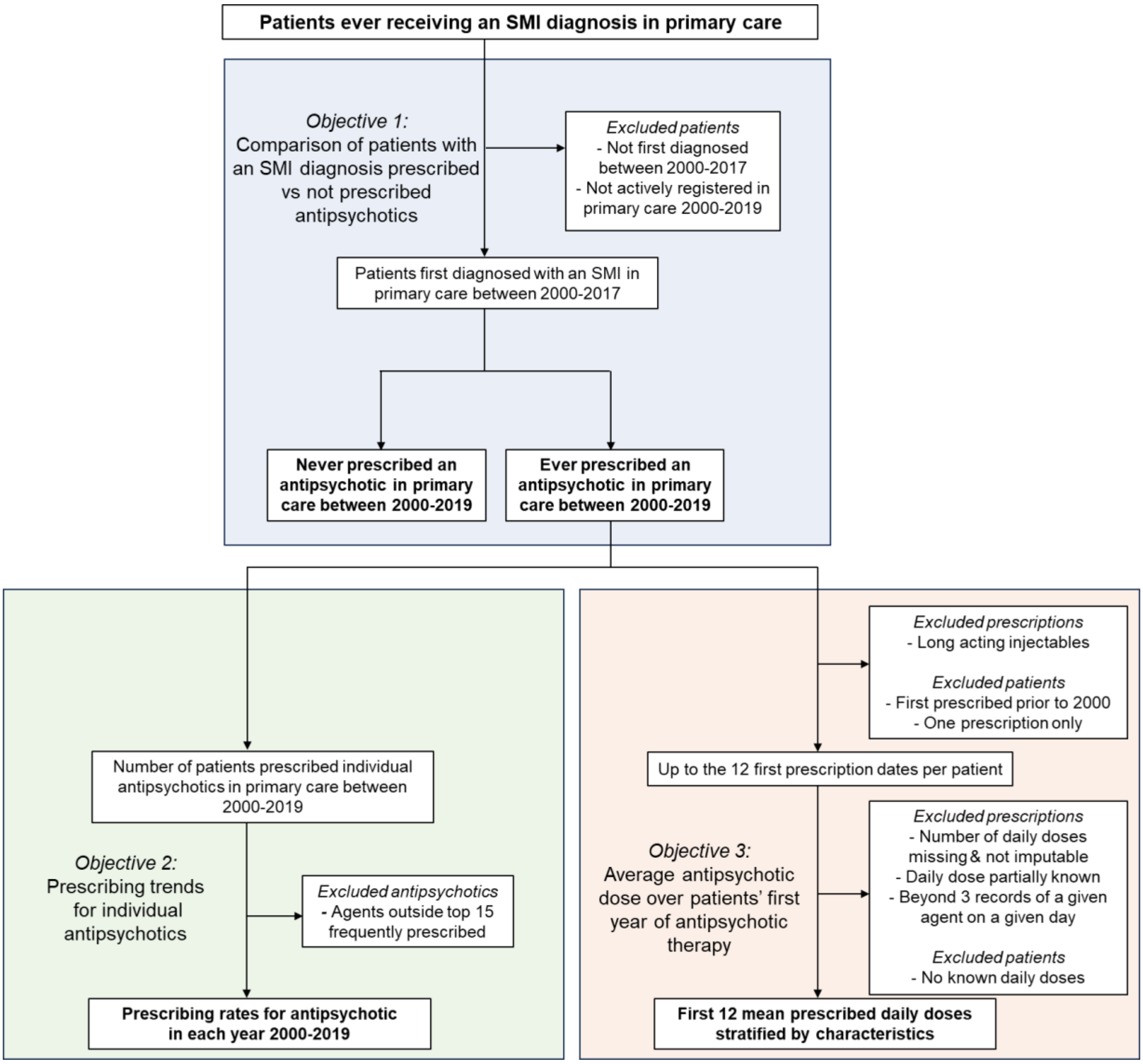

**Supplementary Figure 1. Study design.**

** Boxes in bold represent the main output of each objective.*

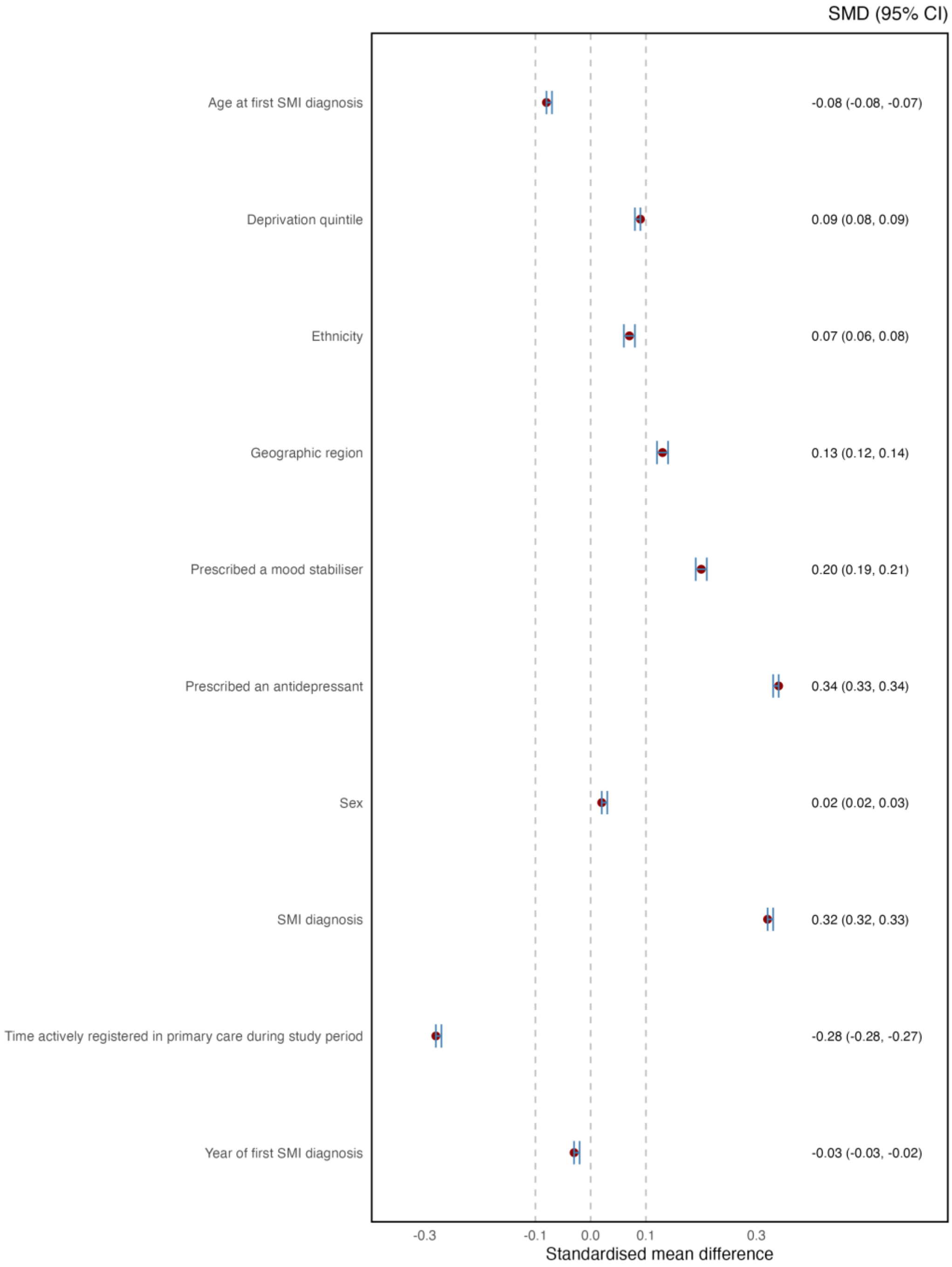

**Supplementary Figure** 2**. Forest plot of standardised mean differences in the comparison of patients prescribed and not prescribed antipsychotics.**

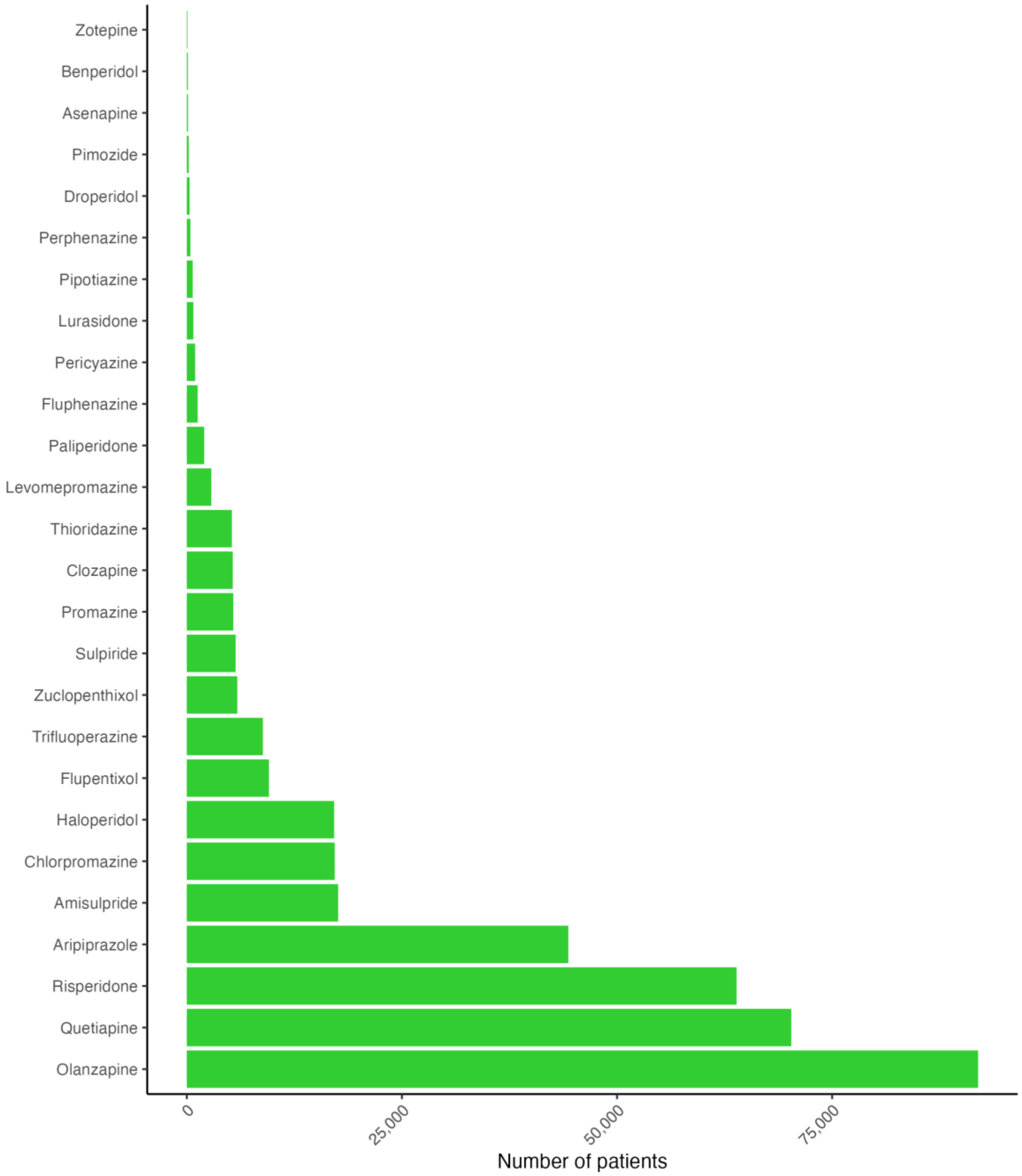

**Supplementary Figure 3. Number of patients prescribed each antipsychotic.**

*The overall total number of patients in the cohort prescribed at least one antipsychotic between 2000-2019 was 212,618. Individual patients are counted more than once if prescribed more than one antipsychotic between 2000-2019. Antipsychotics are shown if prescribed to at least 50 patients over the study period.*

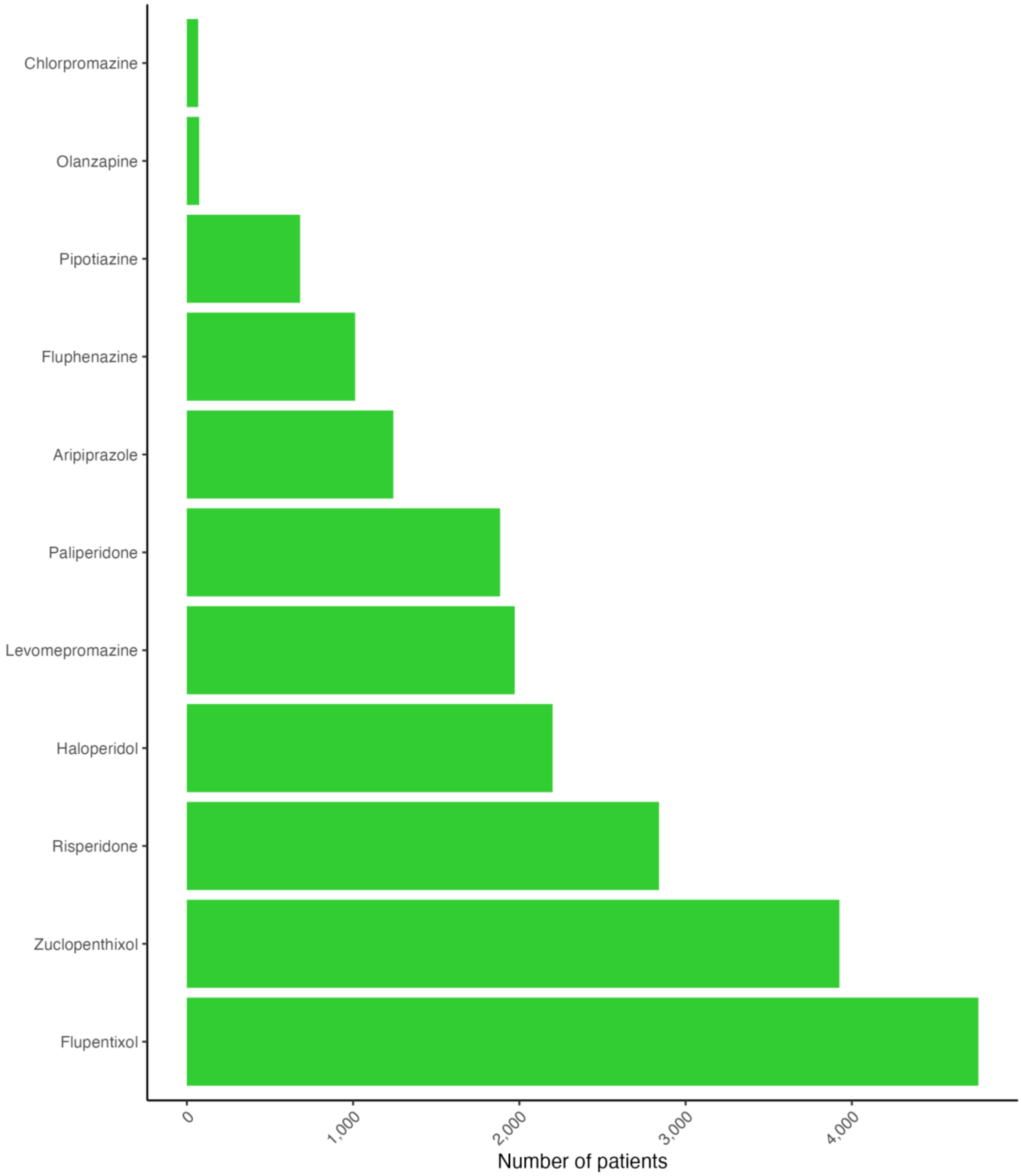

**Supplementary Figure 4. Number of patients prescribed each long-acting injectable antipsychotic.**

*A total of 17,976 patients were identified as ever prescribed a long-acting injectable between 2000-2019. Individual patients are counted more than once if prescribed more than one antipsychotic between 2000 and 2019. Antipsychotics are shown if prescribed to at least 50 patients over the study period.*

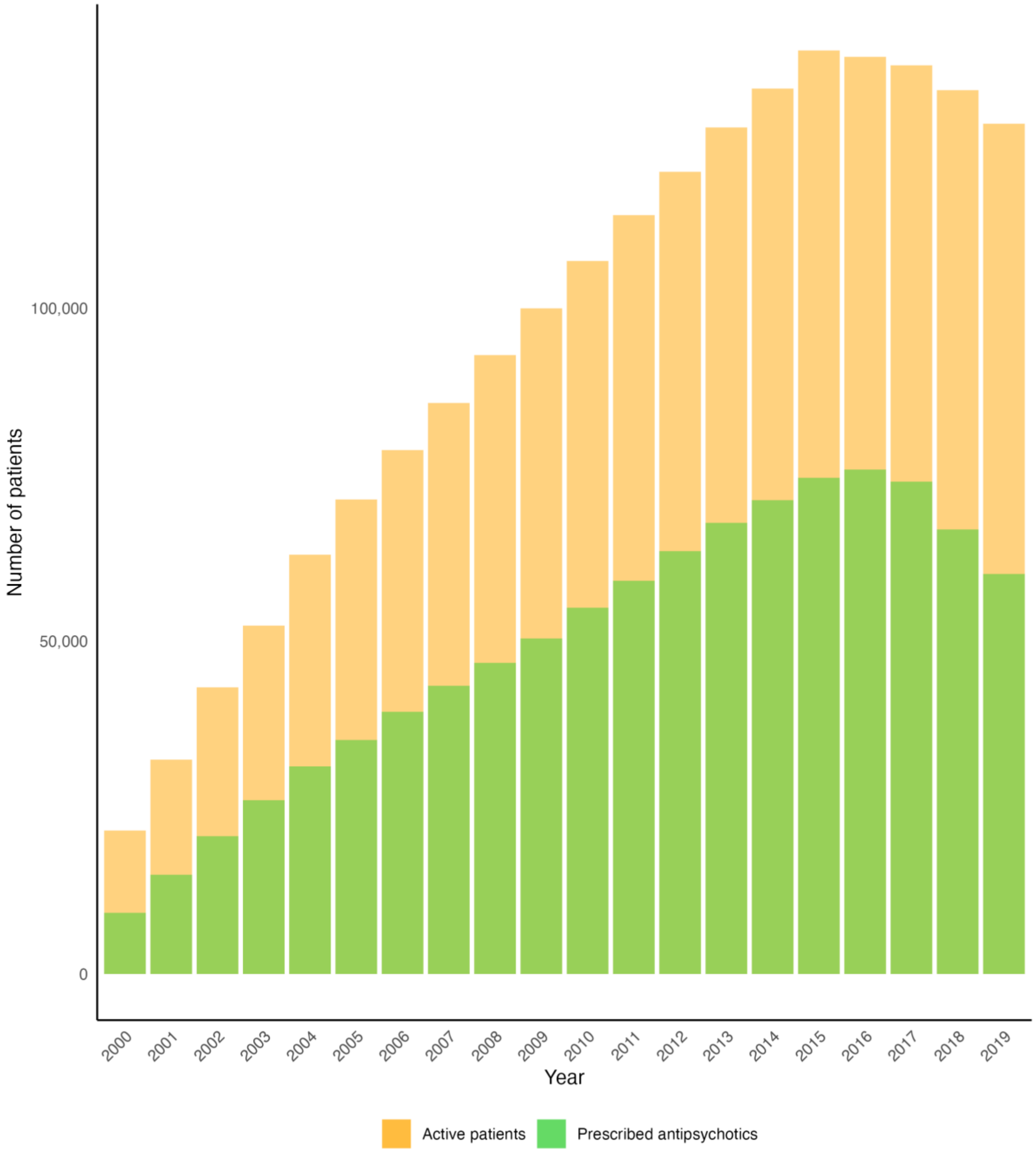

**Supplementary Figure 5. Proportion of patients prescribed antipsychotics 2000-2019.**

*The overall prevalence of antipsychotic prescribing was 426 (95% CI, 420 to 433) per 1,000 patients in the year 2000, reaching a peak of 550 (95% CI, 547 to 553) in 2016, then decreasing to 470 (95% CI, 468 to 473) in 2019.*

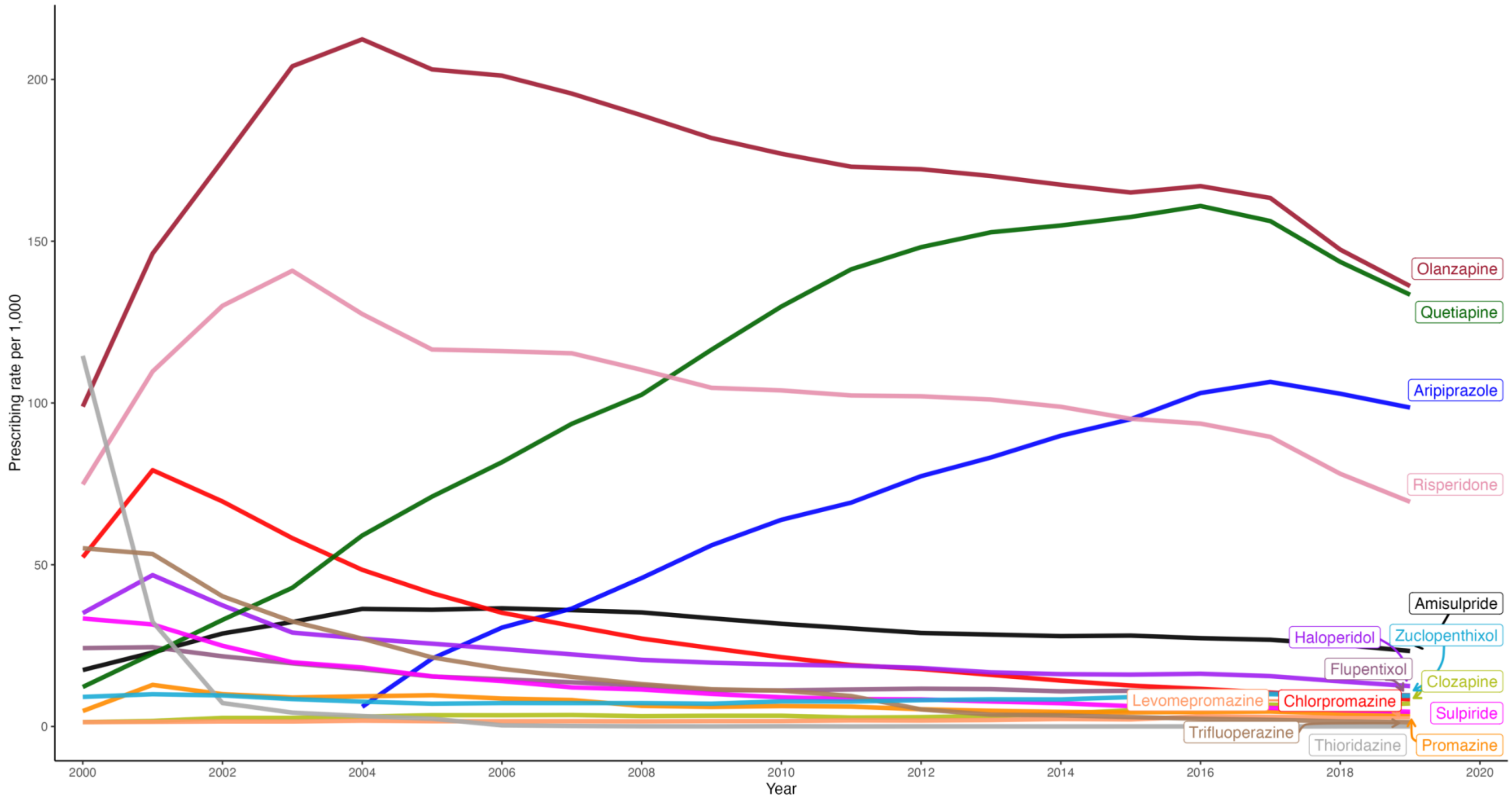

**Supplementary Figure 6. Annual prevalence rates for the prescribing of antipsychotics to patients diagnosed with a severe mental illness (i.e., schizophrenia, bipolar disorder, and other non-organic psychoses).**

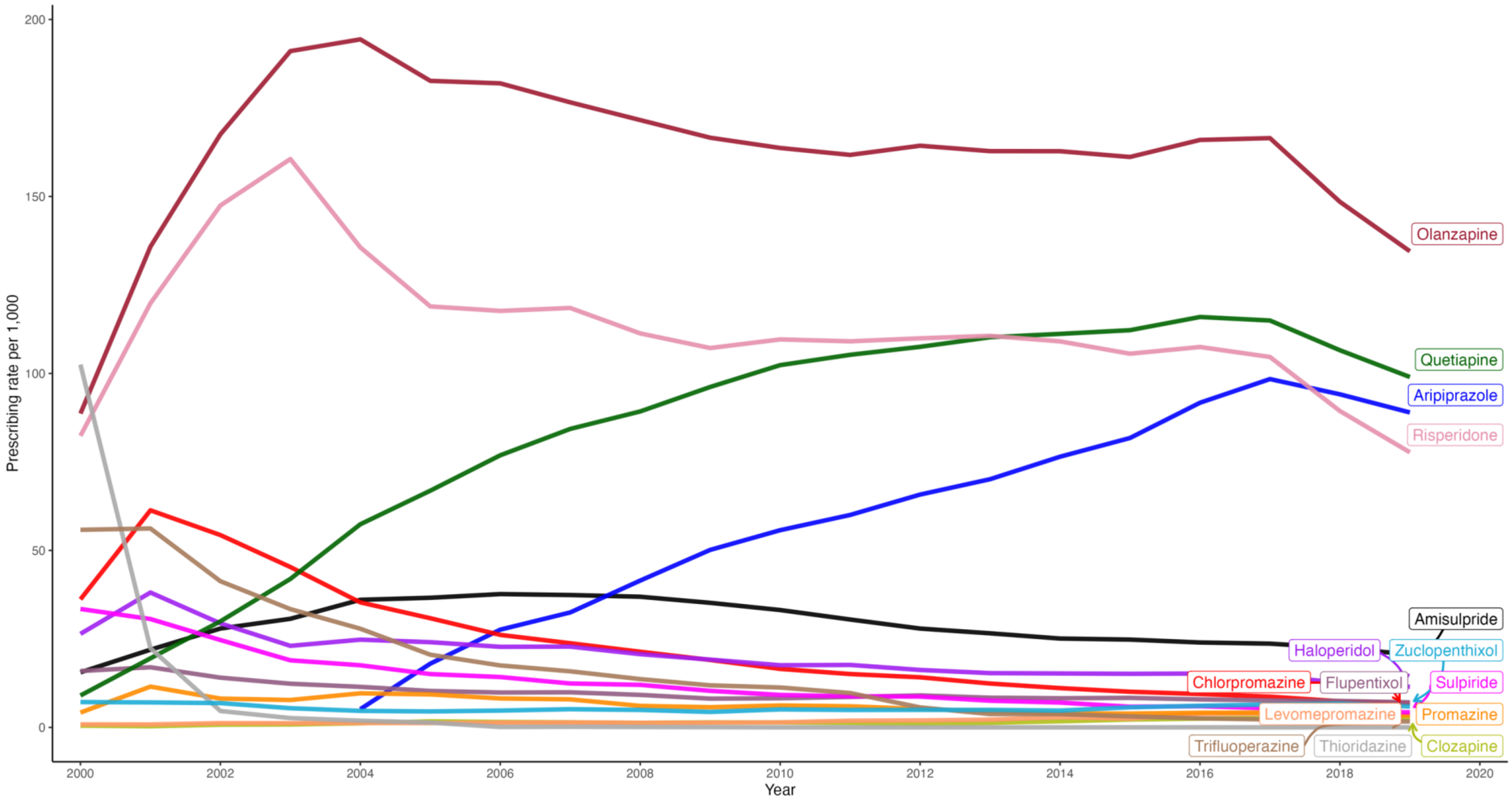

**Supplementary Figure 7. Annual prevalence rates for the prescribing of antipsychotics to patients diagnosed with other non-organic psychoses.***

** Other non-organic psychoses included diagnoses such as psychotic episodes, schizoaffective disorders, delusional disorder, and non-organic psychosis not otherwise specified.*

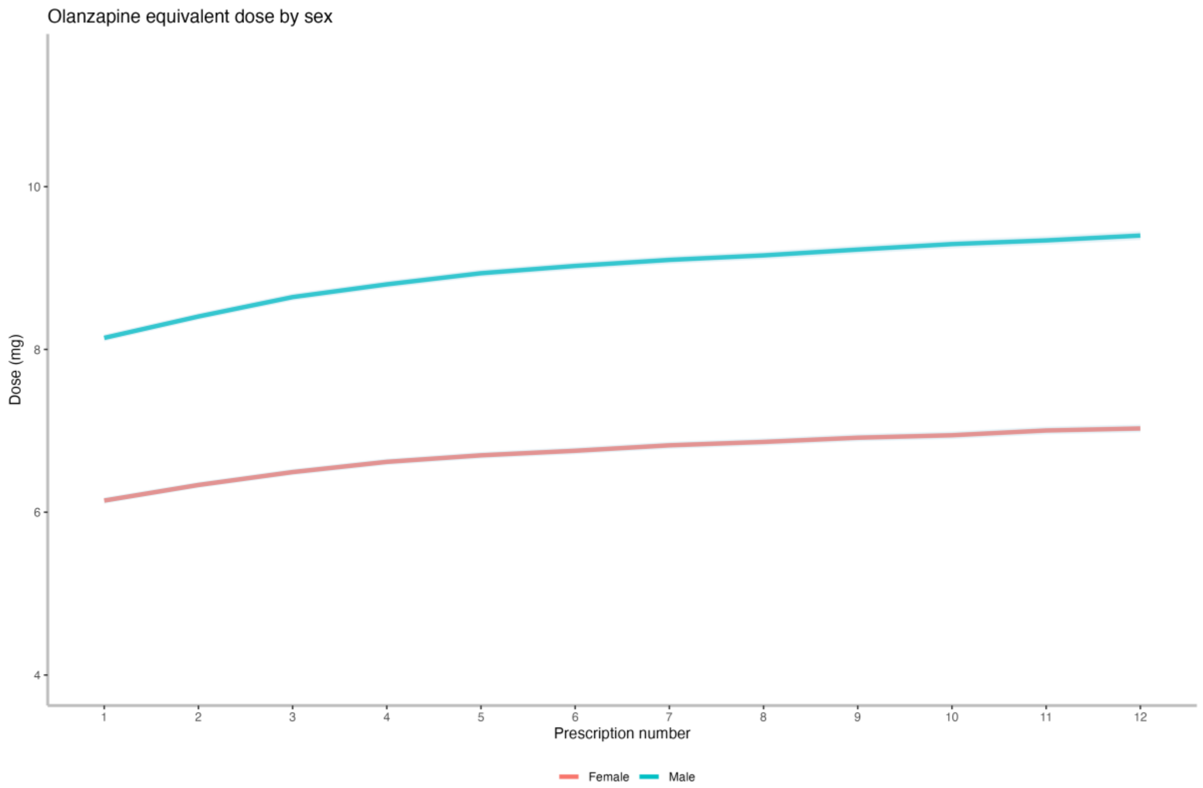

| Group | Dose 1 | Dose 6 | Dose 12 |
|---|---|---|---|
| Female | 6.1 (5.8) [n=86,036] | 6.8 (5.9) [n=71,966] | 7.0 (6.1) [n=59,914] |
| Male | 8.1 (6.8) [n=93,104] | 9.0 (6.8) [n=75,907] | 9.4 (7.0) [n=62,153] |

**Supplementary Figure 8. Mean total daily prescribed oral antipsychotic dose over the first 12 prescriptions – stratified by sex.**

*The graph shows the mean olanzapine equivalent dose (mg) at each time-point, with 95% confidence intervals. The table beneath the graph shows the corresponding mean (SD) olanzapine equivalent doses at prescription date 1, 6 and 12, with the number of observations at each time-point. The overall median time between prescription dates was 28 days.*

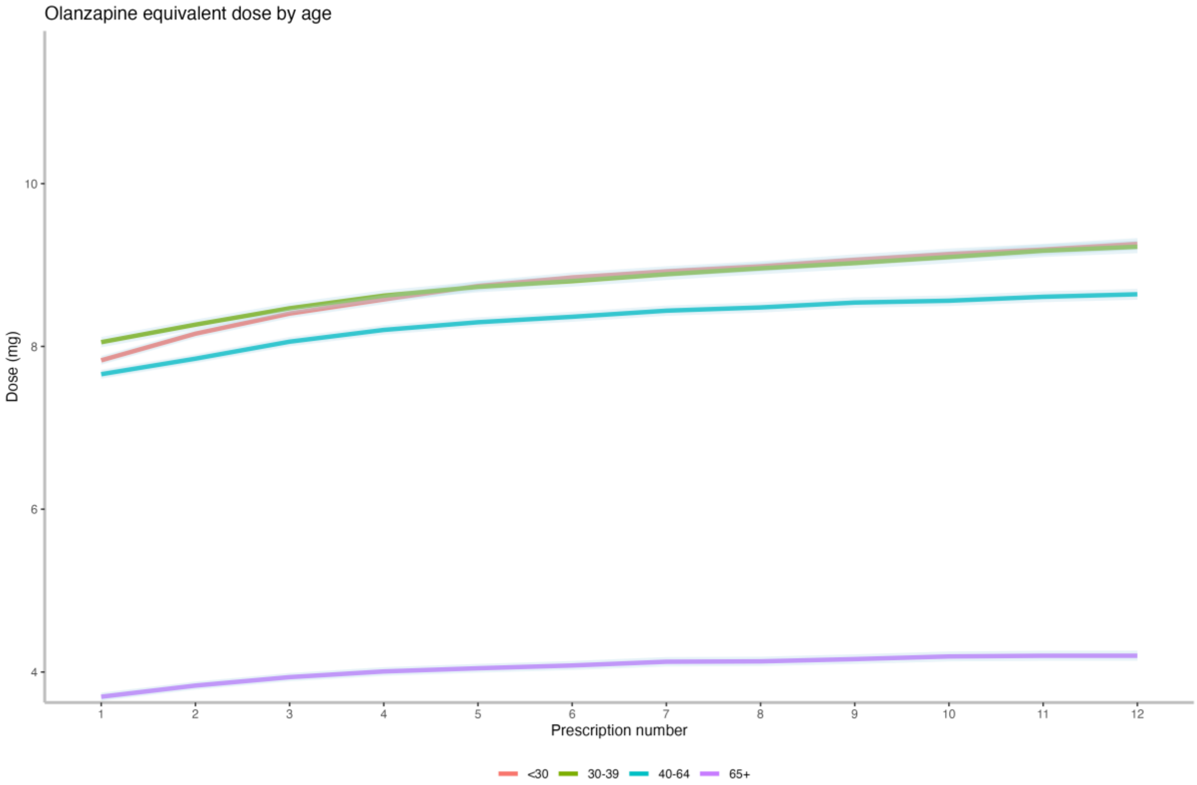

| Group | Dose 1 | Dose 6 | Dose 12 |
|---|---|---|---|
| <30 | 7.8 (6.3) [n=53,044] | 8.8 (6.4) [n=41,851] | 9.3 (6.6) [n=32,913] |
| 30-39 | 8.1 (6.6) [n=42,658] | 8.8 (6.5) [n=35,121] | 9.2 (6.8) [n=29,007] |
| 40-64 | 7.7 (6.6) [n=55,370] | 8.4 (6.8) [n=47,290] | 8.6 (6.8) [n=40,630] |
| 65+ | 3.7 (4.3) [n=28,079] | 4.1 (4.3) [n=23,619] | 4.2 (4.4) [n=19,522] |

**Supplementary Figure 9. Mean total daily prescribed oral antipsychotic dose over the first 12 prescriptions – stratified by age group.**

*The graph shows the mean olanzapine equivalent dose (mg) at each time-point, with 95% confidence intervals. The table beneath the graph shows the corresponding mean (SD) olanzapine equivalent doses at prescription date 1, 6 and 12, with the number of observations at each time-point. The overall median time between prescription dates was 28 days.*

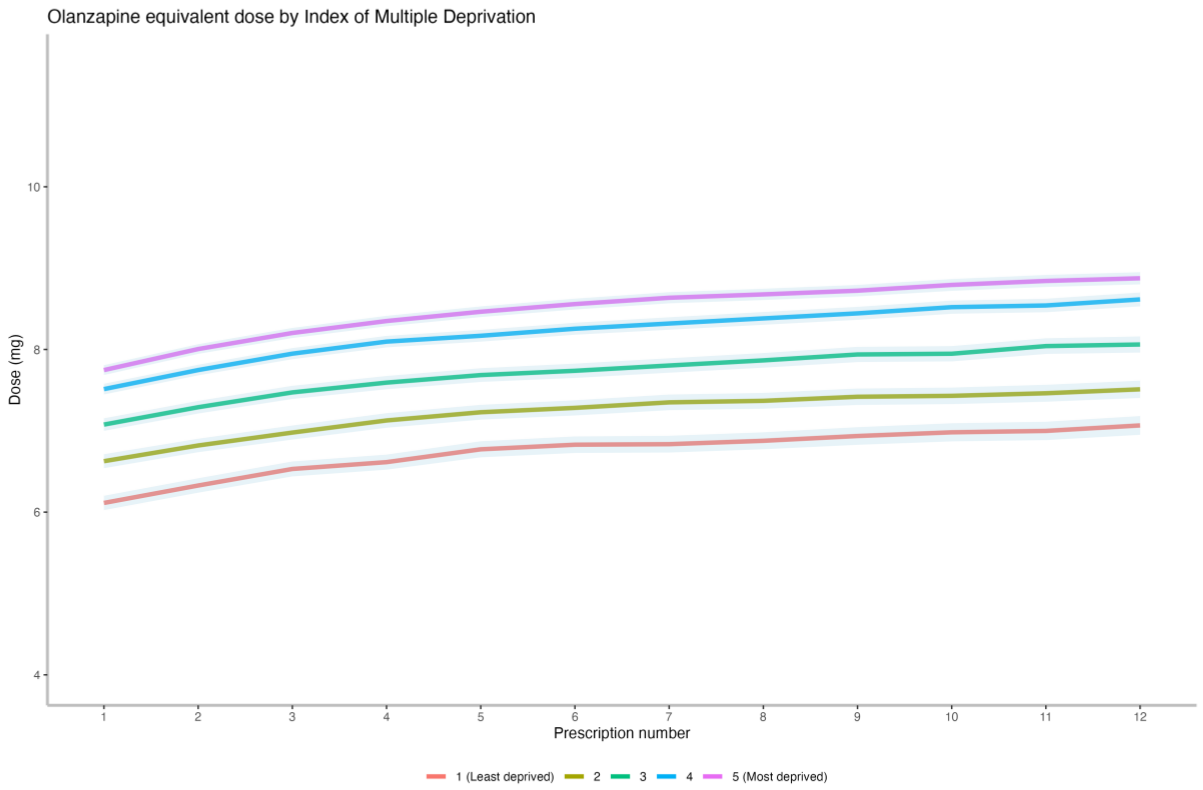

| Group | Dose 1 | Dose 6 | Dose 12 |
|---|---|---|---|
| 1 (Least deprived) | 6.1 (5.8) [n=17,092] | 6.8 (6.1) [n=13,817] | 7.1 (6.2) [n=11,212] |
| 2 | 6.6 (6.3) [n=21,049] | 7.3 (6.2) [n=17,150] | 7.5 (6.4) [n=13,979] |
| 3 | 7.1 (6.4) [n=26,806] | 7.7 (6.6) [n=21,962] | 8.1 (6.8) [n=17,926] |
| 4 | 7.5 (6.4) [n=36,910] | 8.3 (6.5) [n=30,193] | 8.6 (6.8) [n=24,871] |
| 5 (Most deprived) | 7.7 (6.6) [n=46,055] | 8.6 (6.6) [n=38,637] | 8.9 (6.8) [n=32,401] |

**Supplementary Figure 10. Mean total daily prescribed oral antipsychotic dose over the first 12 prescriptions – stratified by quintile of the 2019 English Index of Multiple Deprivation.**

*The graph shows the mean olanzapine equivalent dose (mg) at each time-point, with 95% confidence intervals. The table beneath the graph shows the corresponding mean (SD) olanzapine equivalent doses at prescription date 1, 6 and 12, with the number of observations at each time-point. The overall median time between prescription dates was 28 days.*
